# Integrating dynamic nomogram and machine learning for personalized disability prediction in elderly cardiometabolic multimorbidity: routine blood markers and mental health

**DOI:** 10.64898/2026.06.25.26356637

**Authors:** Xiaojin Huang, Shuo Yang, Lianqing Ma, Tonghui Song, Jiaqi Li, Xingyu Zhang, Haowen Xue, Shuchang Cao, Wenbo Yan, Sihang Zhang, Shuqin Sun

## Abstract

**Background:** Disability prediction in elderly with cardiometabolic multimorbidity (CMM) is limited. We developed a dynamic nomogram and addressed three questions: predictive value of routine blood markers, depression vs. physical function, and plateau in CMM count.

**Methods:** Using CHARLS data (46 predictors), disability defined as ADL/IADL impairment or self-report. LASSO and logistic regression built the nomogram, with mediation, RCS, trend tests, machine learning, and SHAP.

**Results:** Six predictors (depression, cognition, stroke, CMM number, age, falls) formed a good-performing nomogram (https://xjbsashjtdx.shinyapps.io/DynamicNomogram/). Left-hand grip strength mediated 12.3% of stroke’s effect. Cognition showed an inverted U-shape (inflection point=12.043). CMM count plateaued after 3 diseases. Depression outranked grip strength and walking speed. SHAP identified HbA1c, creatinine, uric acid, hematocrit, fasting glucose, TyG, and CVAI as risk markers.

**Conclusions:** The nomogram enables personalized risk stratification. Routine blood markers predict disability, depression dominates over physical function, and the CMM-disability relationship plateaus at CMM≥3.

## Introduction

The World Health Organization (WHO) defines chronic diseases as conditions of prolonged duration and typically slow progression, not transmissible between individuals, primarily including cardiovascular diseases (CVD), diabetes, and chronic respiratory diseases [1]. According to China’s Sixth National Health Service Survey Report, the prevalence of chronic diseases rises with age, reaching 62.3% among individuals aged ≥65 years in 2018 [2]. Chronic diseases have become the leading threat to physical health worldwide [3], causing over 41 million deaths globally in 2017, accounting for 71.3% of total deaths [4]. By 2019, the disease burden attributable to chronic diseases exceeded 70% of the global total.

The global population aged ≥65 years reached 703 million in 2019 (13.2% of the global population) and is projected to reach 1.5 billion by 2050 [5]. In China, this trend is particularly pronounced, with the elderly population reaching 190.64 million in 2020 (13.5% of the national population) [6]. Among the elderly, chronic diseases often manifest as significant comorbidities, particularly cardiometabolic multimorbidity (CMM), defined as the simultaneous presence of two or more cardiometabolic diseases in an individual [7, 8]. CMM is associated with adverse long-term prognosis; epidemiological evidence indicates a two-fold increased mortality risk and 12–15 years of reduced life expectancy compared to those with a single cardiometabolic disease [9]. Moreover, the risk of disability is significantly higher in elderly CMM patients than in non-CMM patients [10, 11]. Disability, in turn, leads to falls, hospitalizations, and mortality [12–14], diminishing patients’ self-care ability and quality of life while imposing substantial economic burdens on healthcare systems, families, and society [15–19]. Therefore, early identification of high-risk individuals is crucial for preventing disability and its associated adverse outcomes in elderly CMM patients.

Previous studies have identified various disability-related variables, including age, gender, body mass index (BMI), social activities, alcohol consumption, smoking, depression symptoms, cognitive function, gait speed, and grip strength [20–22]. Although Activities of Daily Living (ADL) and Instrumental Activities of Daily Living (IADL) scales are commonly used to assess disability severity [23], they provide only general assessments and lack individualized risk predictions. Dynamic nomograms enable precise personalized risk prediction [24, 25], while machine learning offers novel approaches for risk modeling in complex diseases through nonlinear fitting and high-dimensional data processing [26, 27].

Currently, clinical prediction models for disability risk in elderly patients with CMM remain limited. To address this gap, we used the China Health and Retirement Longitudinal Study (CHARLS) to identify predictive variables and develop an online dynamic nomogram. Additionally, by integrating traditional regression, mediation analysis, nonlinear analysis, linear trend assessment, and machine learning, this study addresses three key questions: (1) whether routine blood-based metabolic markers predict disability; (2) whether depression symptoms outweigh grip strength and walking speed; and (3) whether the number of CMM exhibits a plateau rather than a purely linear trend. We aim to provide a practical tool for early risk stratification and to guide clinical strategies, including mental health assessment and routine blood testing, for elderly CMM patients.

## Methods

### Statement

The China Health and Retirement Longitudinal Study (CHARLS) is a publicly accessible database. The original ethics approval documents, their English translations, and the exact date when data were retrieved for the present research are unavailable. Researchers may apply for data access via the official CHARLS website(http://charls.pku.edu.cn/en).Throughout the period of data extraction and after the completion of this study, the authors had no access to any personally identifiable information of participants. All individual-level data released by the CHARLS database have been fully de-identified without identifiers such as full names, identity numbers, detailed residential addresses and exact birth dates, so specific respondents cannot be recognized from the analytical dataset.

### Study population and study design

In this study, CMM was defined as individuals suffering from two or more of the following diseases: hypertension, heart disease, stroke, diabetes, and dyslipidemia, all of which were based on self-reported medical diagnoses. Specifically, heart disease includes myocardial infarction, coronary artery disease, angina pectoris, congestive heart failure, and other cardiac conditions. The WHO defines population aged ≥60 years as elderly1.Consequently, the study population comprises individuals aged ≥60 years with CMM.

We used data from the CHARLS, which is publicly accessible at http://charls.pku.edu.cn/en. The CHARLS survey project was approved by the Biomedical Ethics Committee of Peking University (IRB No.00001052–11015), and all participants were required to sign informed consent. Ethics approval for the use of CHARLS was obtained from the University of Newcastle Human Research Ethics Committee (H-2015-0290). Owing to its high quality and large sample size, the data provide authentic and effective support for the analysis presented in this paper28.

Data from the 2011 and 2015 CHARLS waves were analyzed. Following exclusion of participants with >30% missing data, a total of 1, 424 patients aged ≥60 years with CMM were included in the analysis. These participants were randomly divided into a training set comprising 996 individuals and an internal validation set comprising 428 individuals, in a 7:3 ratio. Additionally, A total of 3, 335 patients aged ≥60 years with CMM from the 2018 and 2020 CHARLS waves were included as the external validation set.The detailed inclusion and exclusion criteria of the study population are presented in Figure 1.

**Figure 1.**
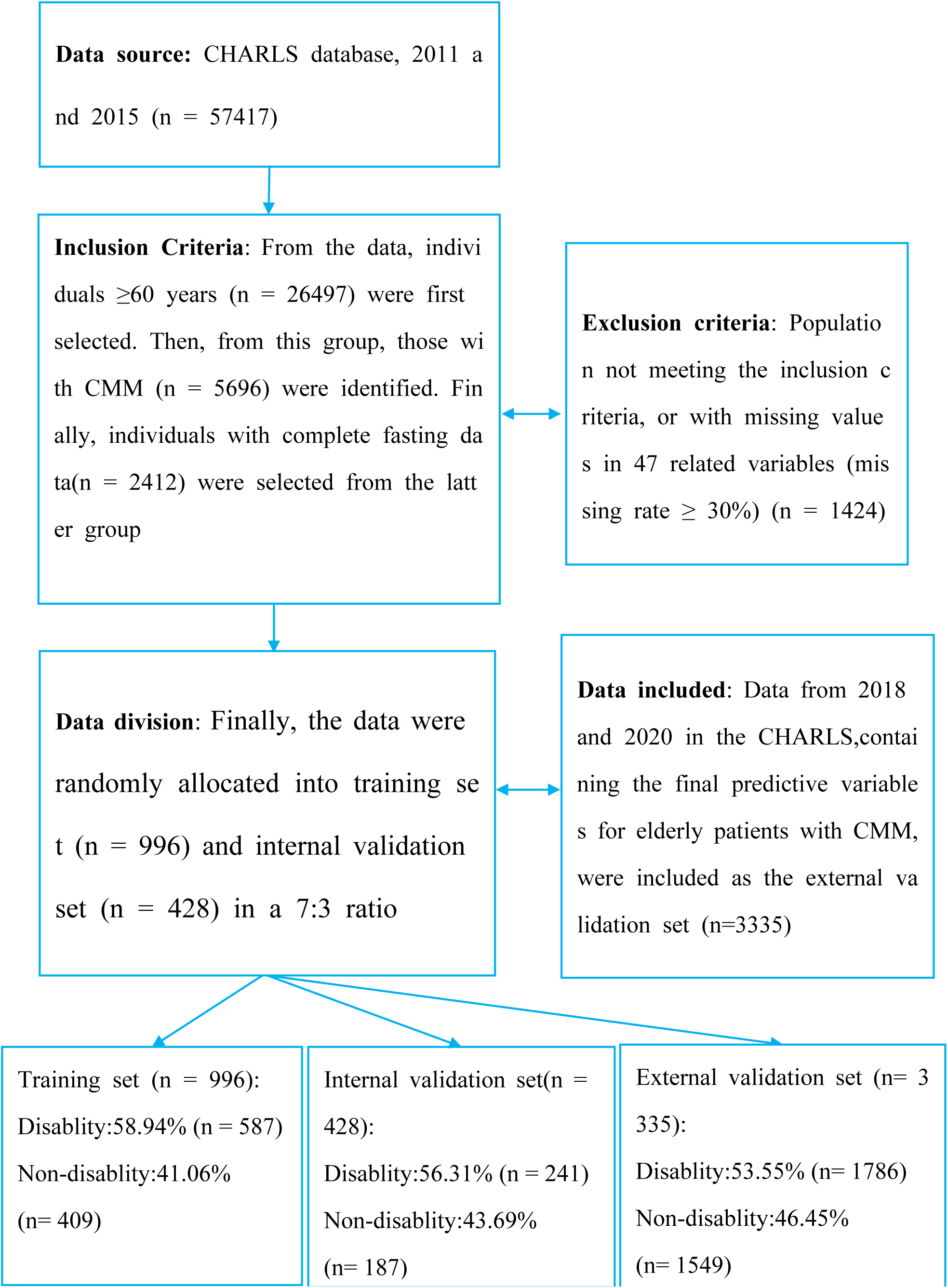
A concise flow chart illustrating the steps for subject selection

### Study variables

#### Outcome variable

The primary outcome variable of interest in this study is disability, defined as limitations in any domain of life due to health or physical issues29.Disability is primarily assessed using ADL and IADL scales30.ADL measures the respondent’s ability to perform basic daily tasks, including dressing, bathing, eating, getting up, using the toilet, and controlling urination and defecation29. IADL assesses the ability to perform more complex instrumental activities, such as doing household chores, preparing hot meals, shopping, managing finances, making phone calls, and taking medication.Participants’ responses were categorized into four levels: (1) no difficulty, (2) difficulty but can still perform the task, (3) difficulty and need assistance, and (4) cannot perform the task. For each ADL/IADL item, a score of 0 was assigned if the participant reported no difficulty, and a score of 1 was assigned if they reported any difficulty or inability to complete the task29. Participants who self-reported one or more disabilities, including physical disability, brain injury or intellectual disability, blindness or partial blindness, deafness or partial deafness, and muteness or severe stuttering, were included. In this study, disability was categorized into two binary outcomes: (1) no disability (ADL and IADL scores = 0 with self-reported non-disabled status) and (2) disability (ADL or IADL score ≥ 1 or self-reported disabled status).

#### Candidate predictive variables

Candidate predictive variables were selected based on clinical relevance, encompassing both established disability-related factors from prior research and novel CMM-associated indicators. Specifically, we incorporated emerging metabolic indices including the Triglyceride Glucose Index (TyG), Triglyceride Glucose-Body Mass Index (TyG-BMI), Chinese Visceral Adiposity Index (CVAI), and the number of CMM.Recent studies have shown that TyG may be a useful tool for identifying high-risk populations for insulin resistance (IR) and diabetes32. TyG is also significantly associated with new-onset cardiovascular disease and outperforms the modified TyG in identifying individuals at risk of cardiovascular events33.Additionally, TyG-BMI is significantly positively correlated with stroke risk and is associated with the severity and short-term outcomes of new-onset acute ischemic stroke. Surprisingly, as TyG-BMI increases, stroke severity decreases, and short-term outcomes improve[34, 35].Moreover, while the traditional obesity index BMI may not fully predict cardiometabolic risk, the CVAI can effectively predict the incidence of metabolic disorders and is linearly related to the risk of CVD, heart disease, and stroke, performing best in predicting CVD events**[36**-3738**39**]. Finally, We are also interested in whether a higher comorbidity count in CMM is associated with an increased risk of disability. In summary, these variables are significantly associated with CMM and may serve as important indicators for predicting the risk of disability in elderly patients with CMM.

Ultimately, a total of 46 factors were identified and selected as candidate predictive variables. These factors are categorized as follows:(1) Demographic and psychological variables: age, gender (male, female), depression symptoms, and life satisfaction.(2) Lifestyle and health behavior variables: sleep duration, social activity, smoking (including are you smoking now and smoking history), and drinking (including are you drinking now and drinking history). (3) Laboratory examination variables: TyG, TyG-BMI, CVAI, glycated hemoglobin A1c (HbA1c), FBG(Fasting Blood Glucose), triglycerides (TG), Total Cholesterol(TC), low-density lipoprotein cholesterol (LDL), high-density lipoprotein cholesterol (HDL), white blood cells (WBC), C-reactive protein (CRP), platelets (PLT), mean corpuscular volume (MCV), hemoglobin (HGB), Hematocrit(HCT), creatinine (CREA), blood urea nitrogen (BUN), cystatin C(CYSC), and uric acid (UA). (4) Physical examination variables: mean of the second and third pulse measurements, maximum left-hand grip strength, maximum right-hand grip strength, average walking speed, BMI, and waist circumference (WC). (5) Clinically relevant variables: chest pain symptoms, cognitive function, fall down, hip fractures, hypertension, heart disease, stroke, diabetes, kidney disease, dyslipidemia, and number of CMM.Among these variables, the binary variables include gender, smoking, drinking, fall down, chest pain symptoms, hip fractures, hypertension, heart disease, stroke, diabetes, kidney disease, and dyslipidemia. The remaining variables are continuous.

### Data collection

#### Demographic and psychological variables, lifestyle and health behavior variables, and clinically relevant variables

The three categories of variables were obtained through questionnaires administered by trained research personnel. Depression symptoms were assessed using the 10-item Center for Epidemiological Studies Depression Scale (CES-D-10), with a total score ranging from 0 to 30, where higher scores indicate more severe depression symptoms40.Life satisfaction was measured on a scale of 1 to 5, with higher scores indicating greater satisfaction. Social activity was quantified by the total number of social activities attended by the participants, with a range of 0 to 1141. These activities included: (1) visiting friends and socializing, (2) playing mahjong, chess, cards, or attending community activity rooms, (3) providing help to relatives, friends, or neighbors not living with the participant, (4) dancing, exercising, or practicing qigong, (5) participating in club activities, (6) volunteer or charity work, (7) caring for individuals with illnesses or disabilities not living with the participant, (8) attending school or training courses, (9) stock trading, (10) internet use, and (11) other social activities. Cognitive function was evaluated by assessing mental status and episodic memory. Mental status was based on components of the telephone interview for cognitive status, which included time orientation, numerical ability, and visual and spatial skills. The mental status score was the sum of correct answers, ranging from 0 to 11. Episodic memory was measured by the average score of immediate and delayed recall of 10 Chinese words, with one point awarded for each correct word recalled, resulting in a total score ranging from 0 to 10. The overall cognitive function score was the sum of the mental status and episodic memory scores, ranging from 0 to 21, with higher scores indicating better cognitive function42.

#### Laboratory examination variables

Blood sample analysis was conducted in two phases. Initially, a complete bloodcell count was performed immediately after sample collection at the local county health center. Subsequently, the samples were transported back to the research headquarters for analysis of other biomarkers. The calculation formulas for TYG, TYG-BMI, and CVAI are as follows[37, 43]:

TYG:

TyG = ln1/2 × Fasting Blood Glu (mg/dL) × Fasting TG (mg/dL);

TyG-BMI:

BMI = Weight (kg)/Height^2^ (m^2^); TyG-BMI = TyG × BMI;

CVAI:

For males: CVAI = - 267.93 + 0.68 × Age (years) + 0.03 × BMI (kg/m^2^) + 4.00 × WC (cm) + 22.00 × log10 (TG) (mmol/L) - 16.32 × HDL (mmol/L);

For females: CVAI = - 187.32 + 1.71 × Age (years) + 4.23 × BMI (kg/m^2)^ + 1.12 × WC (cm) + 39.76 × log10 (TG)(mmol/L) - 11.66 ×HDL(mmol/L).

#### Physical examination variables

Physical examinations of participants were conducted using professional equipment. The mean pulse rate was calculated from the second and third measurements. Maximum grip strength of both hands was assessed using a dynamometer.Walking speed was measured twice with a stopwatch, and the average value was recorded. BMI was calculated based on height and weight (weight/height²). WC was measured using a flexible tape measure.

#### Statistical analysis

All numerical variables are presented as medians and interquartile ranges (for non-normal distributions), and the Wilcoxon rank-sum test is used for intergroupcomparisons.Categorical variables are expressed as percentages, and the chi-square test or Fisher’s exact test is employed for intergroup comparisons. In this study, three methods were used to establish a dynamic nomogram for disability riskin elderly patients with CMM and to conduct internal and external validation.Odds ratios (OR) and 95% confidence intervals (CI) are used as effect estimates.

Initially, 16 related variables were preliminarily selected based on the results of univariate logistic regression in the training set data. Subsequently, 9 significantly related variables were further screened out through Least Absolute Shrinkage and Selection Operator(LASSO) regression analysis.Finally, the 9 screened variables were subjected to multivariate logistic regression analysis, and 6 predictive variables with a P-value < 0.05 were includedin the nomogram prediction model. Additionally, a dynamic nomogram was constructed using the “ShinyPredict” package, which can dynamically predict the risk of disability on a website. To validate the nomogram model, internal and external validations were performed. The performance of the prediction model was then assessed using measures of discrimination, accuracy, and clinical validity.The area under the ROC curve (AUC) was used to determine the model’s discrimination ability. The Hosmer-Lemeshow goodness-of-fit test and calibration curves were conducted to assess model calibration. Decision curve analysis (DCA) was used to assess clinical validity.

The mediation effects of Number of CMM, depression symptoms, cognitive function, Stroke, and Fall down on disability were explored. The mediating variables included CVAI, TyG, TyG-BMI, HCT, HbA1c, social activity, sleep duration, maximum right-hand grip strength, maximum left-hand grip strength, and average walking speed. To detect potential nonlinear relationships, restricted cubic spline (RCS) analysis was performed for cognitive function, depression symptoms, Fall down, and Stroke. Subsequently, threshold effect analysis was applied only to those relationships that exhibited nonlinear patterns, in order to identify inflection points.

For the nominal variable Number of CMM (levels 2–5), logistic regression was performed with level 2 as the reference. And pairwise comparisons were adjusted using Tukey’s method. Given the small sample size in the CMM=5 group (n=13) and the resulting imprecise estimates, we conducted a sensitivity analysis by combining CMM=4 and CMM=5 into a single category (CMM≥4). To assess whether the effect of Number of CMM followed a linear dose–response pattern, three complementary trend analyses were performed: (1) a likelihood-ratio test comparing a continuous linear model with the factor model; (2) orthogonal polynomial contrasts to test linear, quadratic, and cubic trends; and (3) the Cochran–Armitage test for trend. R software was used for all analyses in this study. All tests were two-tailed, and P-value < 0.05 was considered statistically significant.

#### machine learning model development and performance evaluation

For the machine learning analyses, we retained the same training (70%) and internal validation (30%) cohort split and the same 46 candidate variables as in the conventional analyses. The Boruta algorithm was applied to the training cohort, and the resulting Confirmed and Tentative variables were used as the final set of predictors for model construction. Separately, LASSO regression with 10-fold cross-validation was performed on the same 46 variables for sensitivity analysis only; the overlap between LASSO-selected and Boruta-derived variables was examined to confirm the importance of the overlapping predictors.

Five machine learning models were then constructed: Logistic Regression (LR) with strong interpretability, Random Forest (RF) capable of capturing nonlinear relationships, Support Vector Machine (SVM) suitable for high-dimensional data, Light Gradient Boosting (LGB) efficient in processing complex interactive features, and eXtreme Gradient Boosting (XGBoost) with excellent predictive stability. Model performance was comprehensively evaluated using AUC, accuracy, sensitivity, specificity, precision, and F1 score. Calibration curves and Brier scores were applied to assess the consistency between predicted probabilities and actual outcomes. DCA was performed to quantify the clinical net benefit. In addition, the SHapley Additive exPlanations(SHAP) method was used to interpret the feature contribution, impact direction, and individualized prediction mechanism of the optimal model at both global and individual levels.

## Results

### Participant characteristics

In this study, 1424 eligible subjects (including 707 males (49.6%) and 717 females (50.4%)) were randomly divided into a training set (*n*=996) and an internal validation set (*n*=428) in a 7:3 ratio. Among the 1424 subjects, 828 (58.1%) were diagnosed with disability, which further confirmed the high incidence of disability in elderly patients with CMM.Table 1 shows significant intergroup differences (P < 0.001) for most clinical variables, and HCT and HbA1c were also significant in laboratory measures.

**Table 1.**
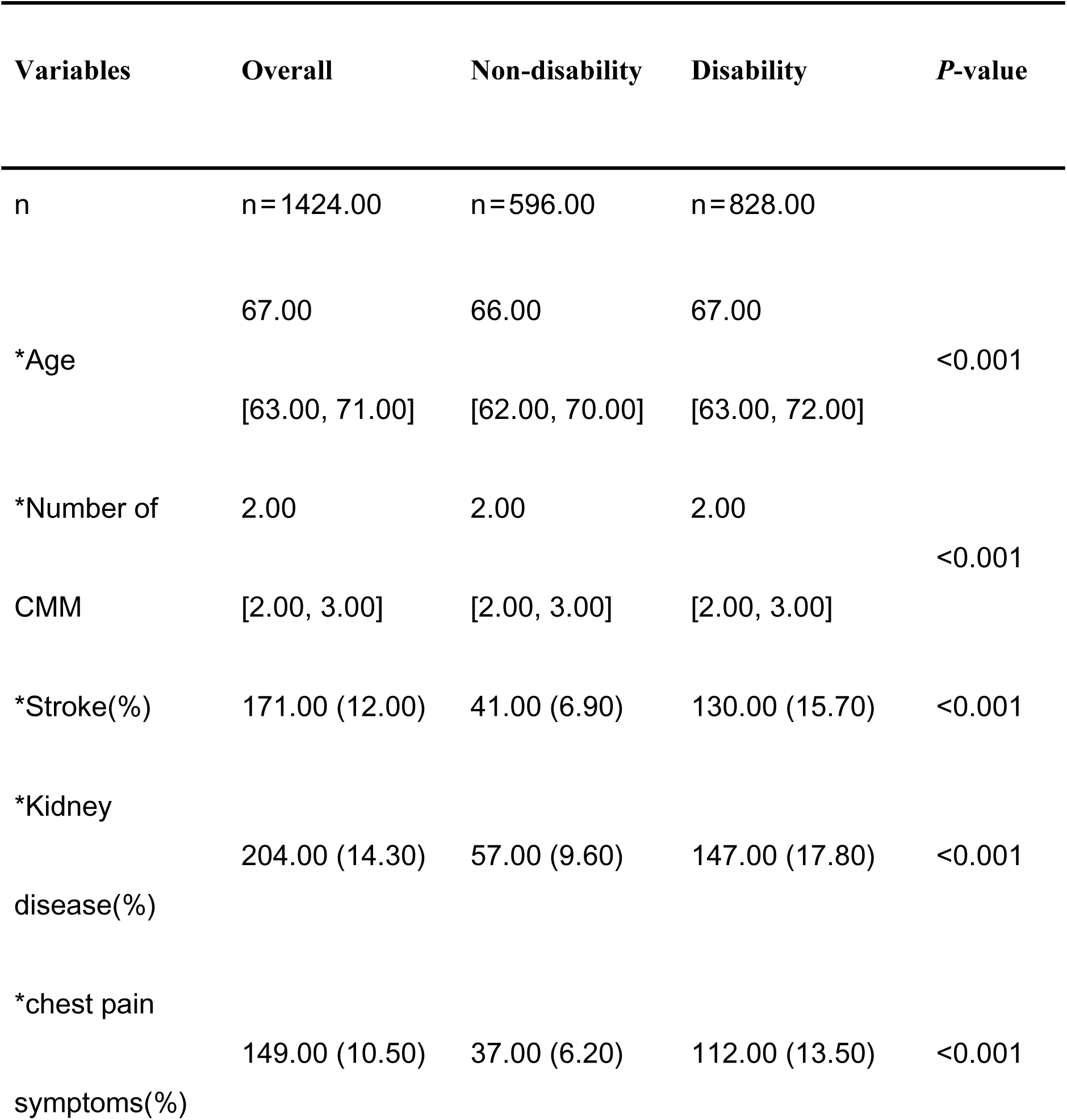
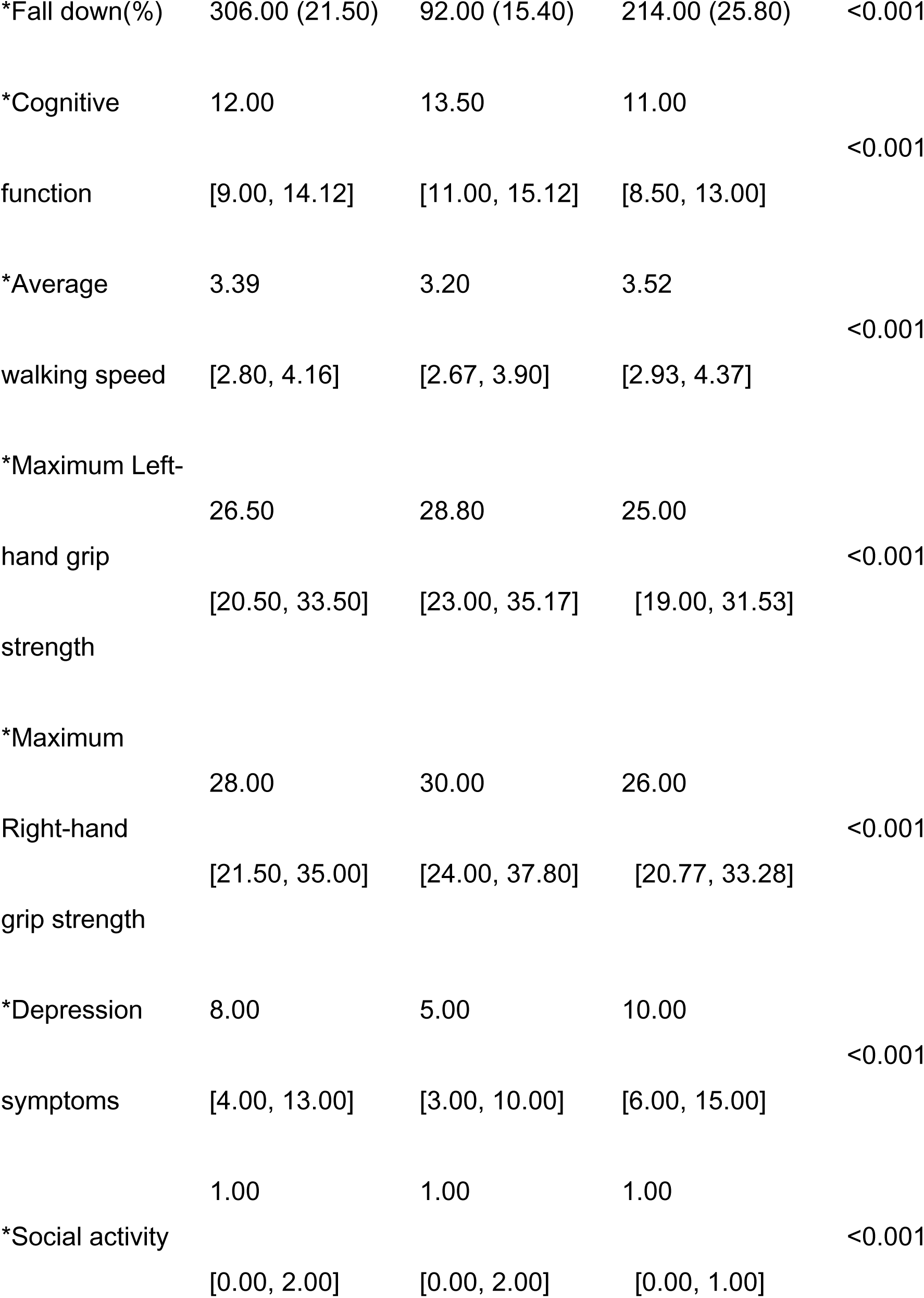
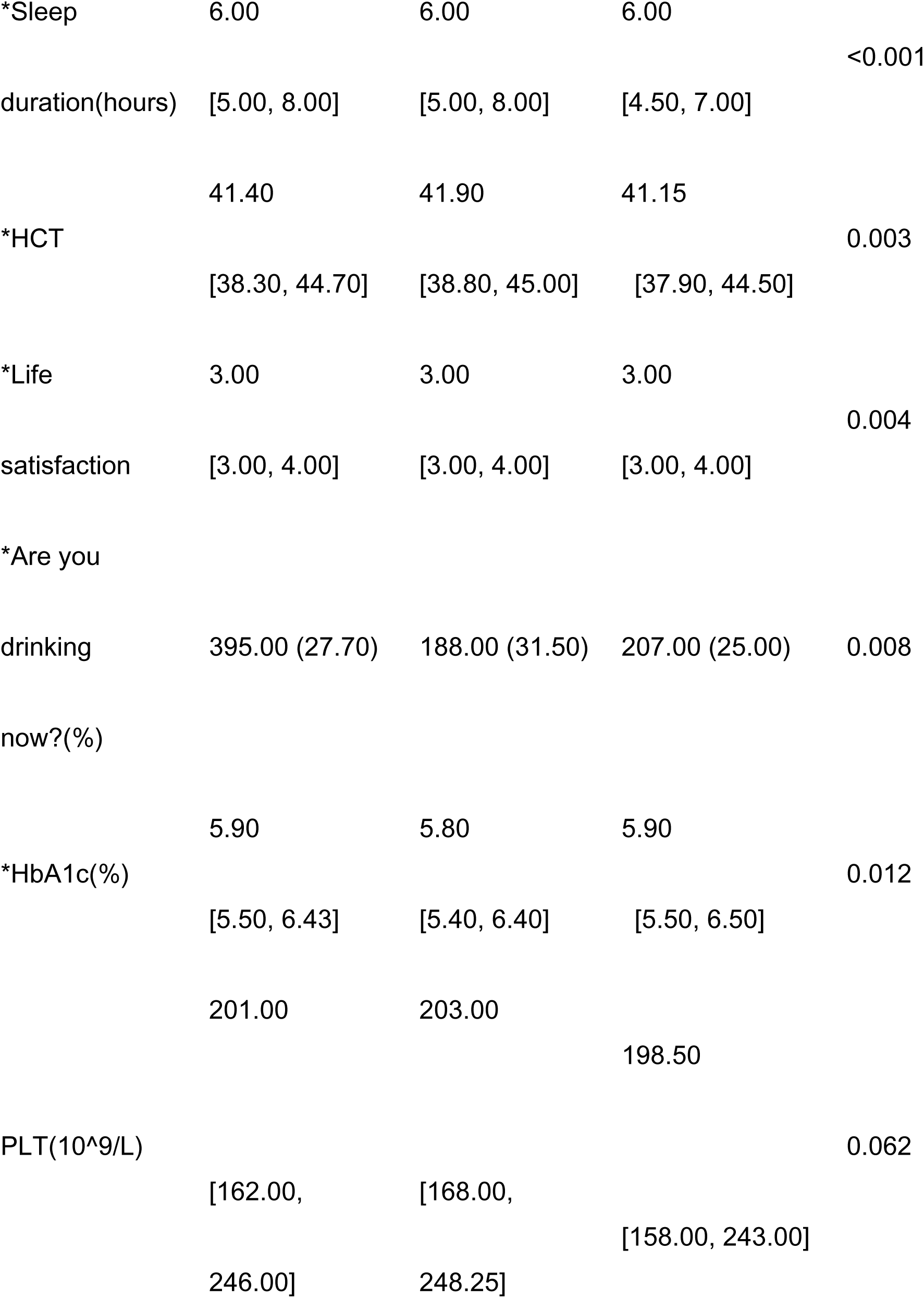
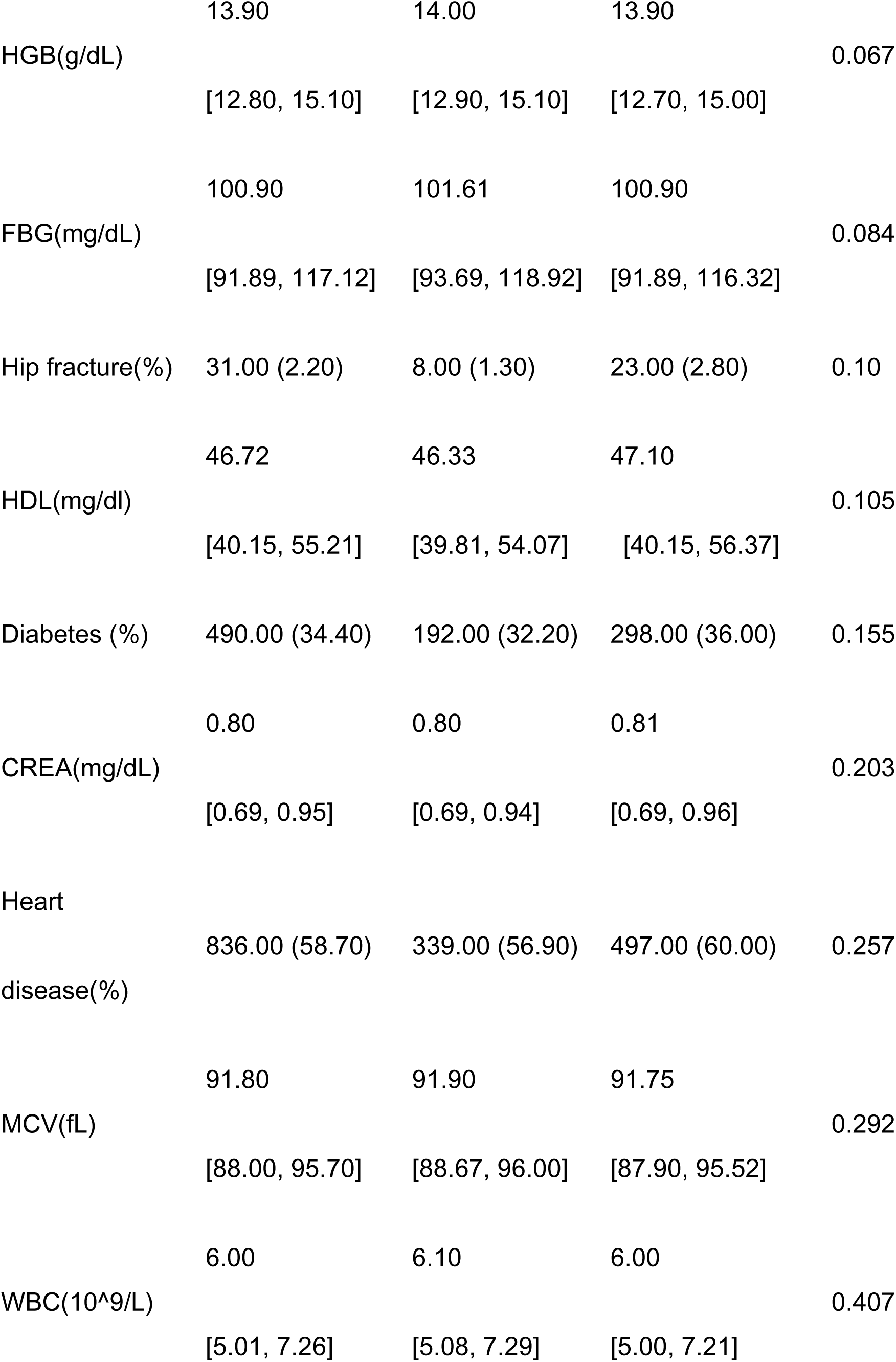
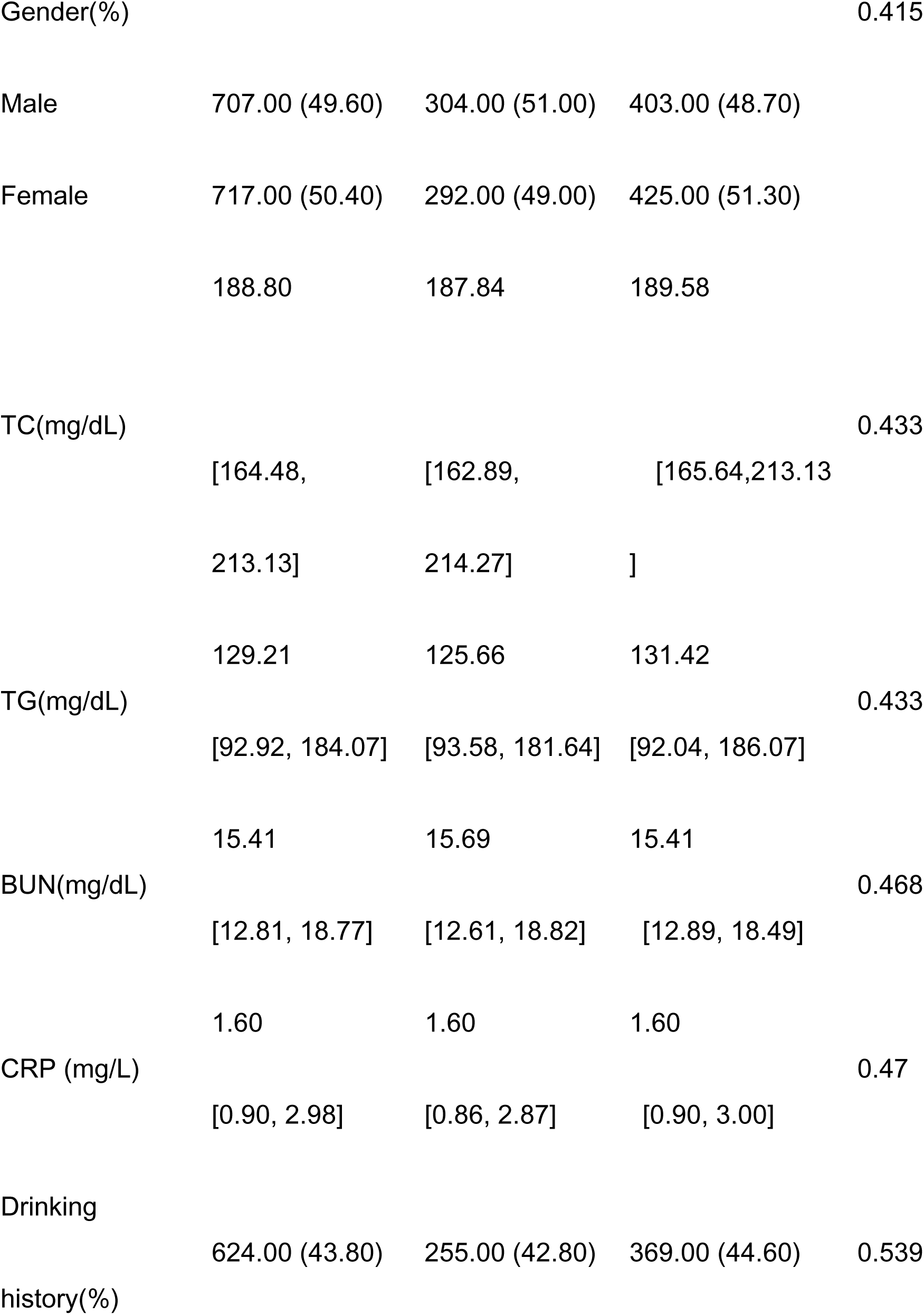
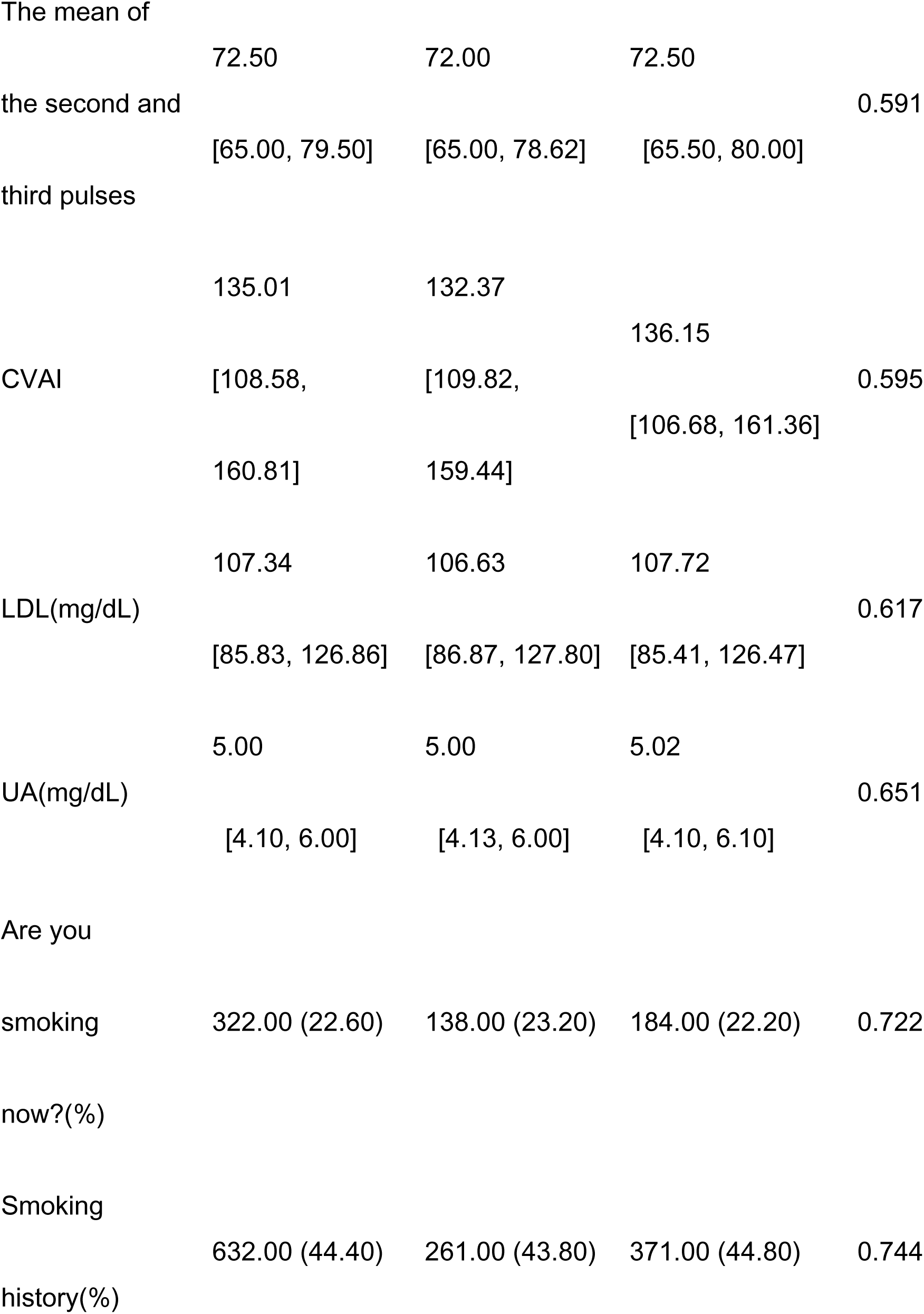
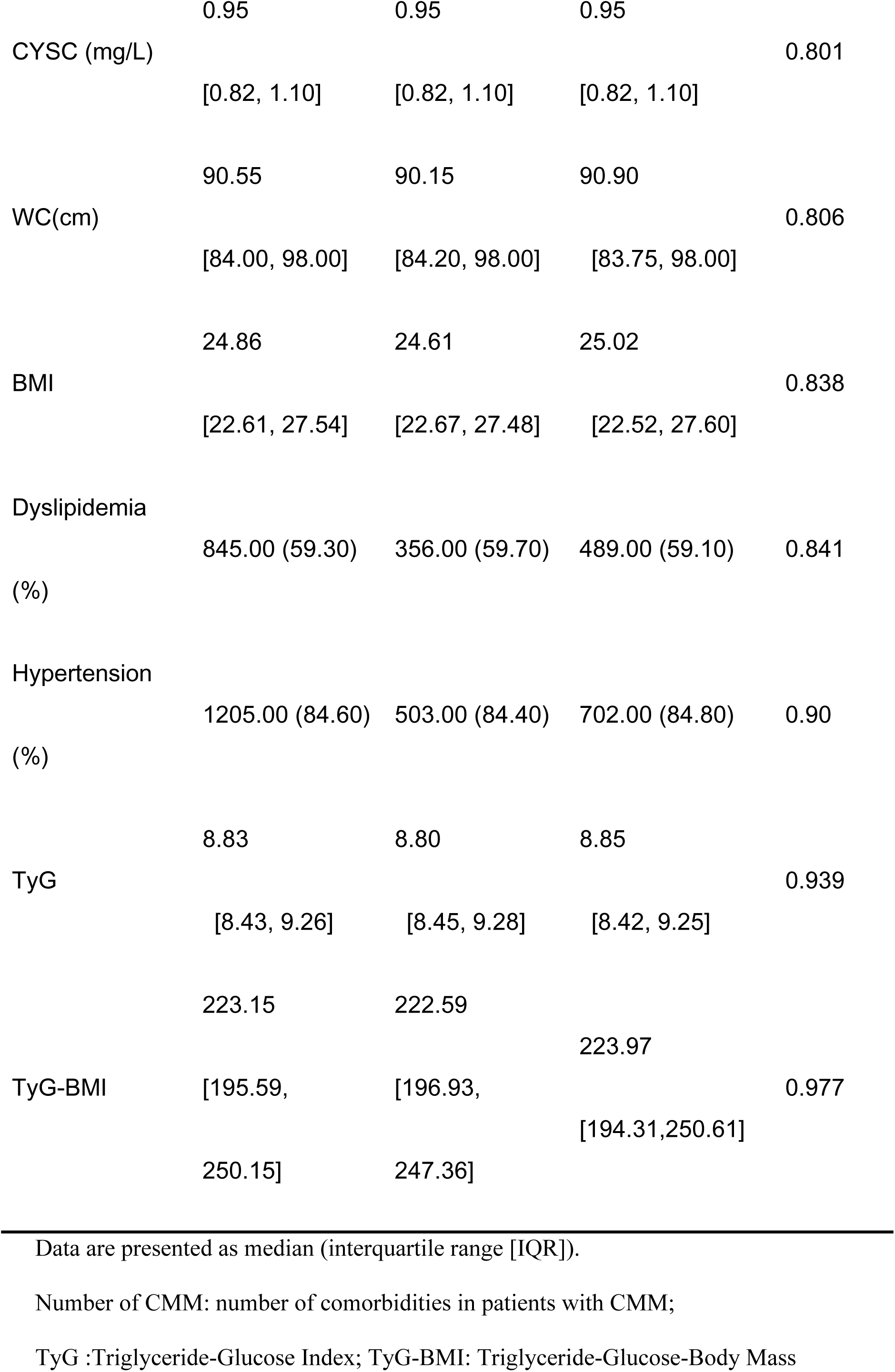
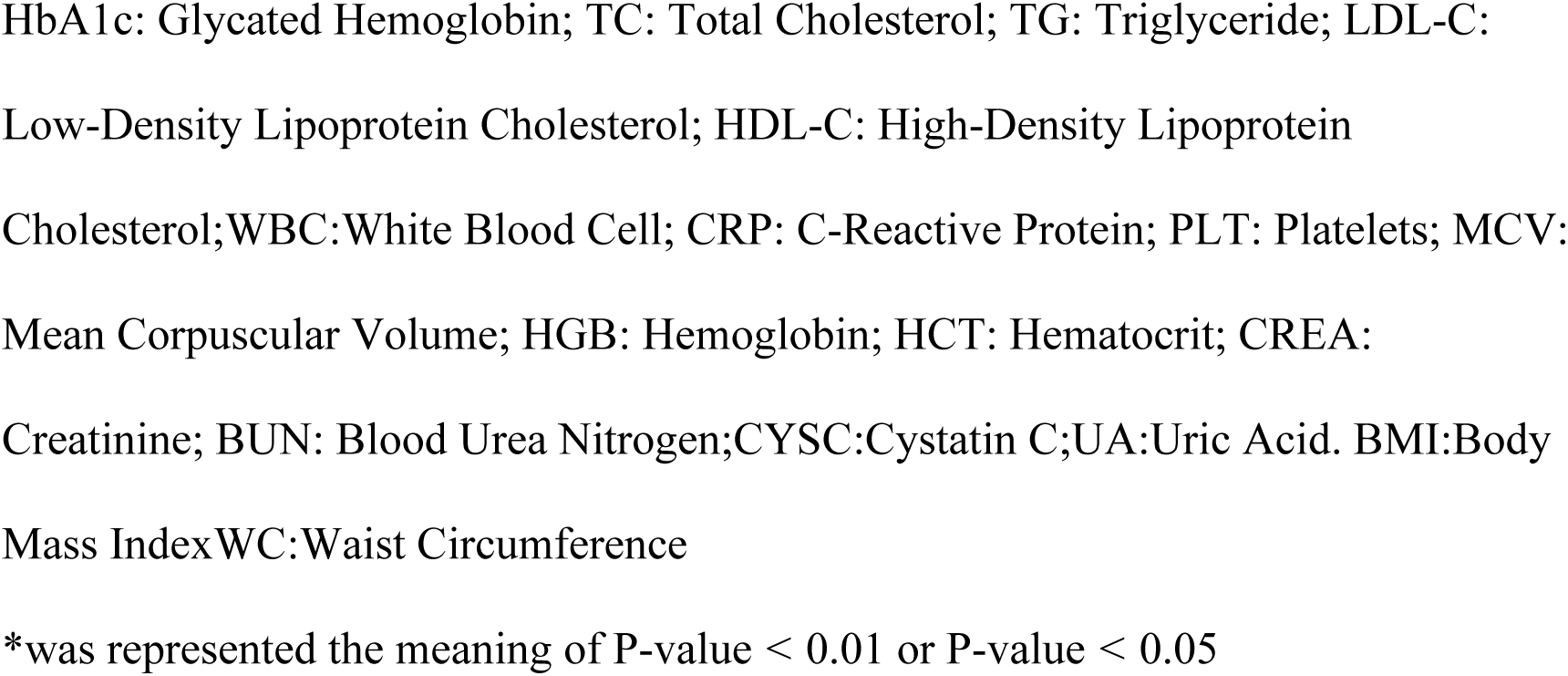
Baseline characteristics of the study population.

### Associations between candidate predictive variables and disability

The results of univariate logistic regression (Tables 2 and 3) showed that age (OR=1.05), number of CMM (OR=1.41), average walking speed (OR=1.24), depression symptoms (OR=1.12), chest pain symptoms (OR=2.43), fall down (OR=2.18), stroke (OR=3.10), kidney disease (OR=1.78), and HDL (OR=1.01) were positively associated with disability (all P<0.05). In contrast, grip strength (both hands, OR=0.95–0.96), sleep duration (OR=0.88), social activity (OR=0.77), cognitive function (OR=0.83), life satisfaction (OR=0.80), and current drinking (OR=0.75) were negatively associated (all P<0.05). Notably, average walking speed and HDL appeared as risk factors, while current drinking was a protective factor.

**Table 2.**
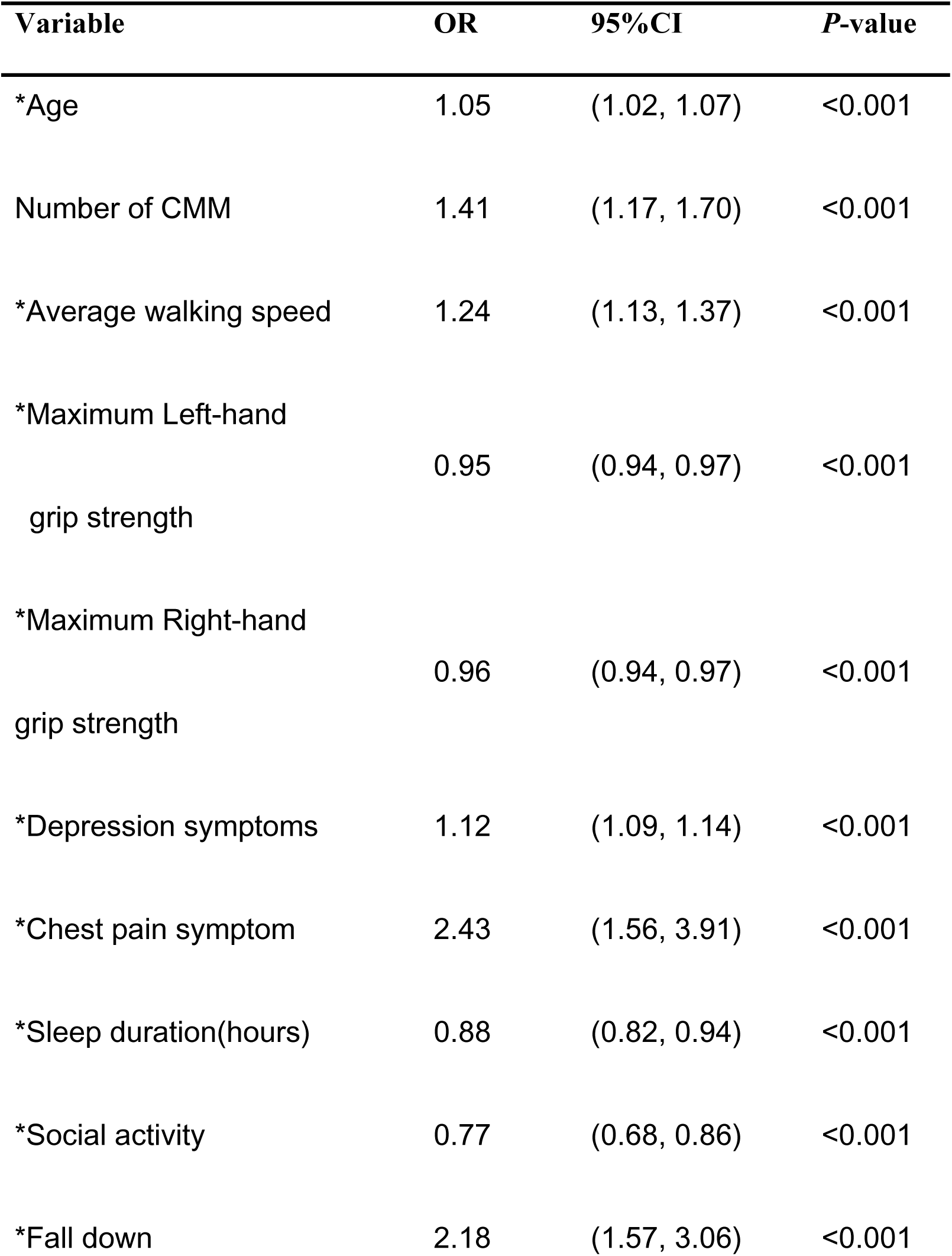
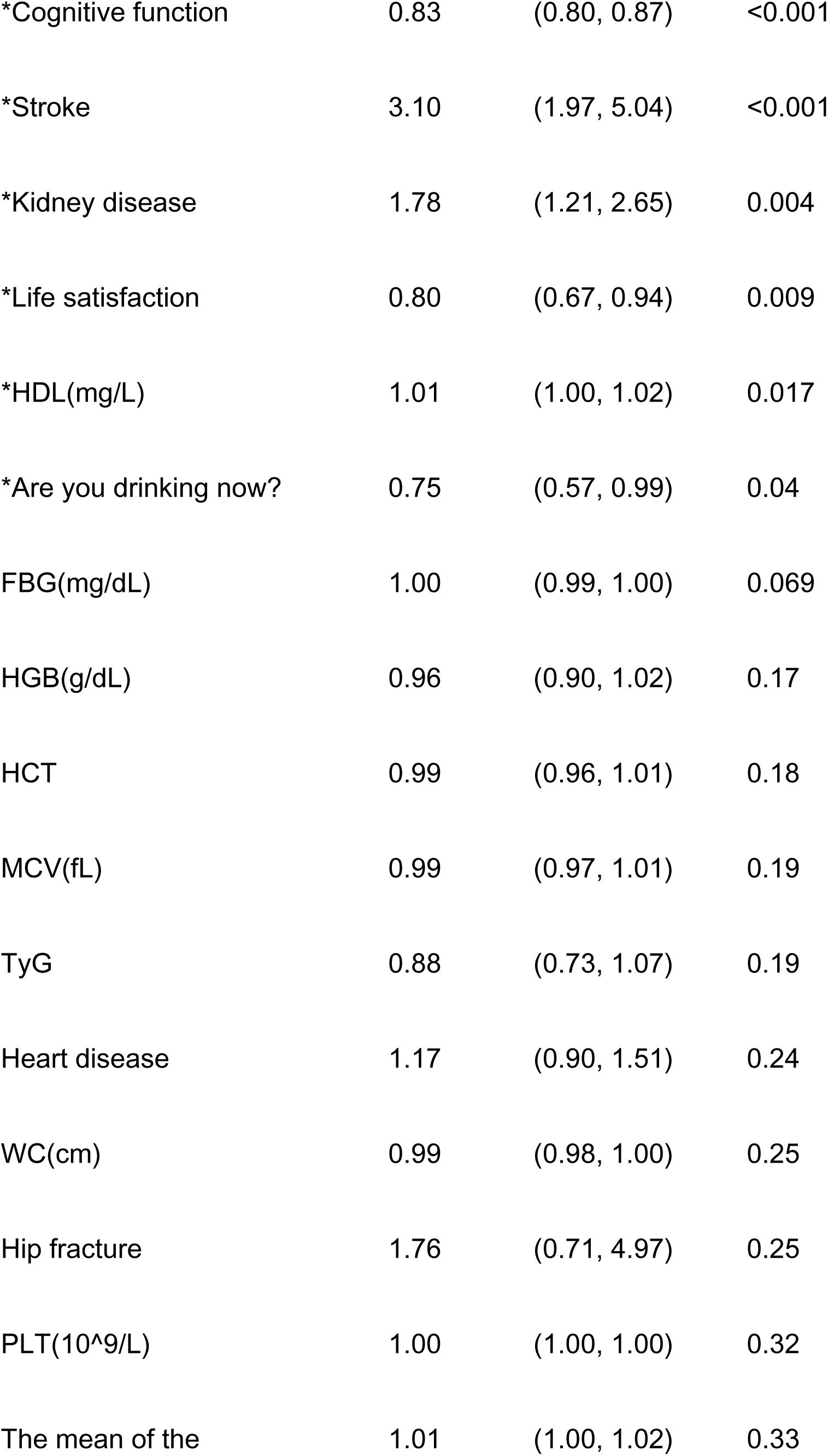
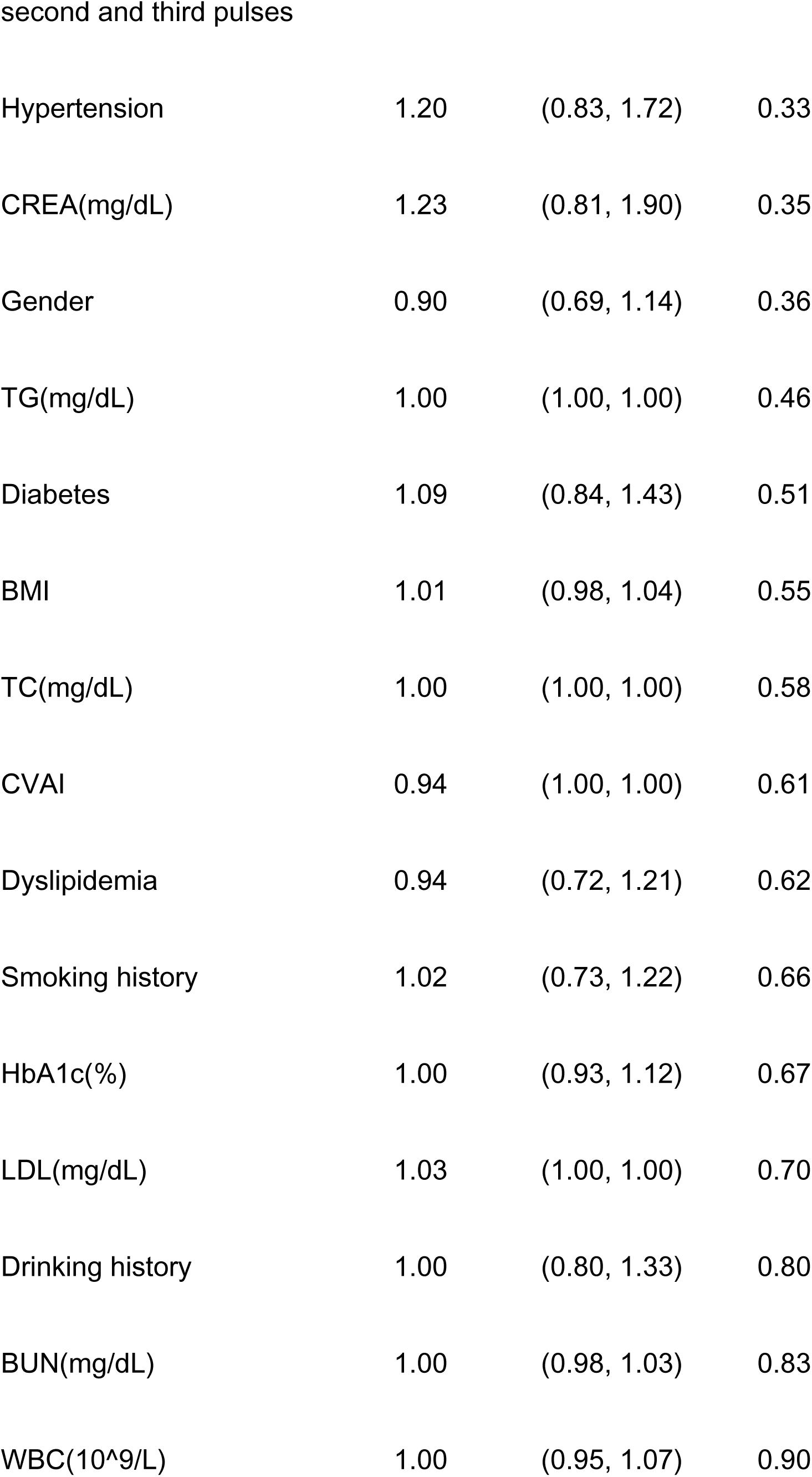
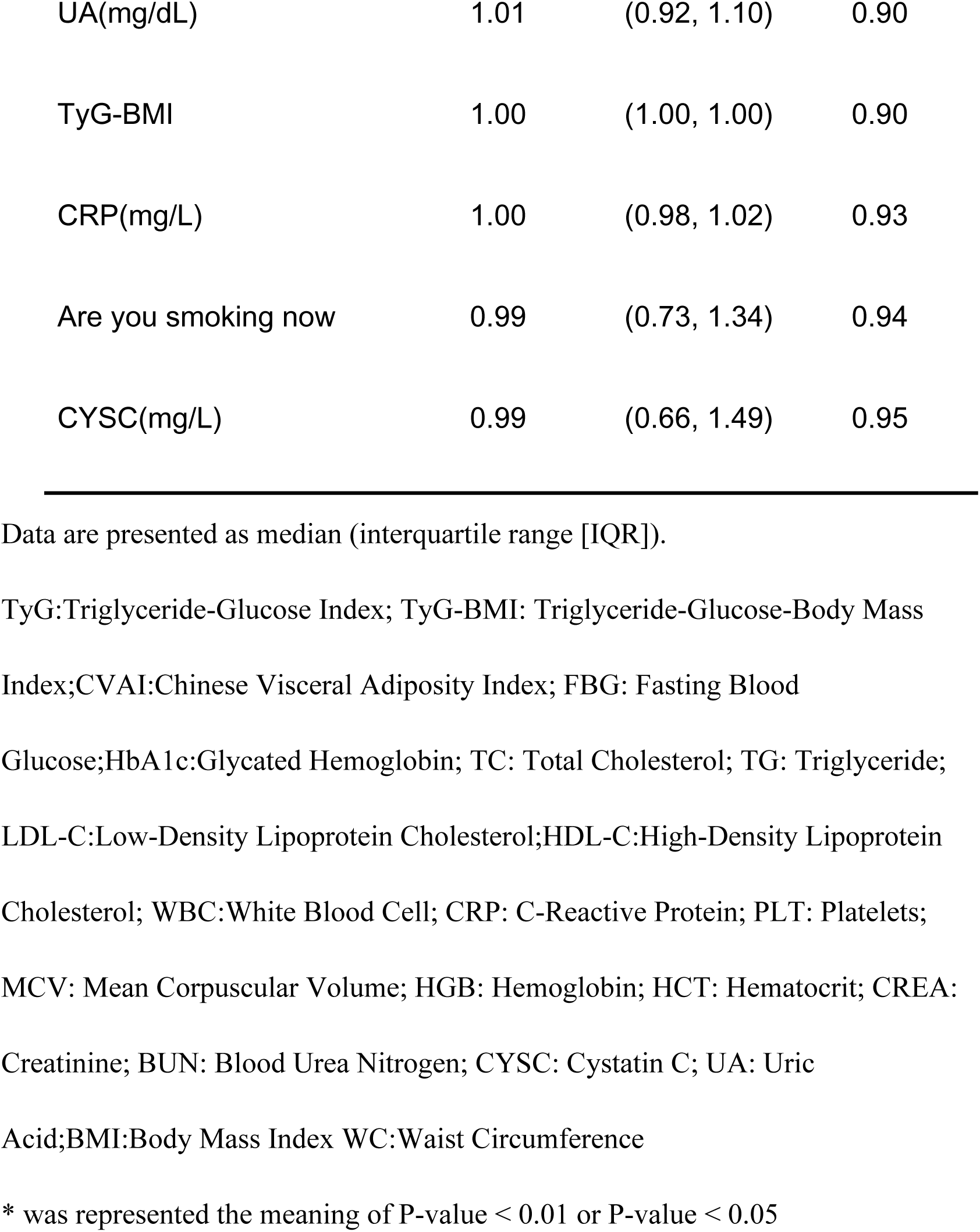
Univariable logistic regression analysis of the Training Cohorts.

| Variable | OR | 95%CI | *P*-value |
| --- | --- | --- | --- |
| *Age | 1.05 | (1.02, 1.07) | <0.001 |
| Number of CMM | 1.41 | (1.17, 1.70) | <0.001 |
| *Average walking speed | 1.24 | (1.13, 1.37) | <0.001 |
| *Maximum Left-hand<br>grip strength | 0.95 | (0.94, 0.97) | <0.001 |
| *Maximum Right-hand<br>grip strength | 0.96 | (0.94, 0.97) | <0.001 |
| *Depression symptoms | 1.12 | (1.09, 1.14) | <0.001 |
| *Chest pain symptom | 2.43 | (1.56, 3.91) | <0.001 |
| *Sleep duration(hours) | 0.88 | (0.82, 0.94) | <0.001 |
| *Social activity | 0.77 | (0.68, 0.86) | <0.001 |
| *Fall down | 2.18 | (1.57, 3.06) | <0.001 |
| *Cognitive function | 0.83 | (0.80, 0.87) | <0.001 |
| *Stroke | 3.10 | (1.97, 5.04) | <0.001 |
| *Kidney disease | 1.78 | (1.21, 2.65) | 0.004 |
| *Life satisfaction | 0.80 | (0.67, 0.94) | 0.009 |
| *HDL(mg/L) | 1.01 | (1.00, 1.02) | 0.017 |
| *Are you drinking now? | 0.75 | (0.57, 0.99) | 0.04 |
| FBG(mg/dL) | 1.00 | (0.99, 1.00) | 0.069 |
| HGB(g/dL) | 0.96 | (0.90, 1.02) | 0.17 |
| HCT | 0.99 | (0.96, 1.01) | 0.18 |
| MCV(fL) | 0.99 | (0.97, 1.01) | 0.19 |
| TyG | 0.88 | (0.73, 1.07) | 0.19 |
| Heart disease | 1.17 | (0.90, 1.51) | 0.24 |
| WC(cm) | 0.99 | (0.98, 1.00) | 0.25 |
| Hip fracture | 1.76 | (0.71, 4.97) | 0.25 |
| PLT($10^9/L$) | 1.00 | (1.00, 1.00) | 0.32 |
| The mean of the | 1.01 | (1.00, 1.02) | 0.33 |
| Hypertension | 1.20 | (0.83, 1.72) | 0.33 |
| CREA(mg/dL) | 1.23 | (0.81, 1.90) | 0.35 |
| Gender | 0.90 | (0.69, 1.14) | 0.36 |
| TG(mg/dL) | 1.00 | (1.00, 1.00) | 0.46 |
| Diabetes | 1.09 | (0.84, 1.43) | 0.51 |
| BMI | 1.01 | (0.98, 1.04) | 0.55 |
| TC(mg/dL) | 1.00 | (1.00, 1.00) | 0.58 |
| CVAI | 0.94 | (1.00, 1.00) | 0.61 |
| Dyslipidemia | 0.94 | (0.72, 1.21) | 0.62 |
| Smoking history | 1.02 | (0.73, 1.22) | 0.66 |
| HbA1c(%) | 1.00 | (0.93, 1.12) | 0.67 |
| LDL(mg/dL) | 1.03 | (1.00, 1.00) | 0.70 |
| Drinking history | 1.00 | (0.80, 1.33) | 0.80 |
| BUN(mg/dL) | 1.00 | (0.98, 1.03) | 0.83 |
| WBC($10^9/L$) | 1.00 | (0.95, 1.07) | 0.90 |
| UA(mg/dL) | 1.01 | (0.92, 1.10) | 0.90 |
| TyG-BMI | 1.00 | (1.00, 1.00) | 0.90 |
| CRP(mg/L) | 1.00 | (0.98, 1.02) | 0.93 |
| Are you smoking now | 0.99 | (0.73, 1.34) | 0.94 |
| CYSC(mg/L) | 0.99 | (0.66, 1.49) | 0.95 |

Data are presented as median (interquartile range [IQR]).
TyG:Triglyceride-Glucose Index; TyG-BMI: Triglyceride-Glucose-Body Mass Index;CVAI:Chinese Visceral Adiposity Index; FBG: Fasting Blood Glucose;HbA1c:Glycated Hemoglobin; TC: Total Cholesterol; TG: Triglyceride; LDL-C:Low-Density Lipoprotein Cholesterol;HDL-C:High-Density Lipoprotein Cholesterol; WBC:White Blood Cell; CRP: C-Reactive Protein; PLT: Platelets; MCV: Mean Corpuscular Volume; HGB: Hemoglobin; HCT: Hematocrit; CREA: Creatinine; BUN: Blood Urea Nitrogen; CYSC: Cystatin C; UA: Uric Acid;BMI:Body Mass Index WC:Waist Circumference
\* was represented the meaning of P-value < 0.01 or P-value < 0.05

**Table 3.** Multivariable logistic regression analysis of the Training Cohorts.

| Characteristic | OR | 95%CI | P-value |
| --- | --- | --- | --- |
| *Age | 1.03 | (1.00,1.06) | 0.021 |
| *Number of CMM | 1.32 | (1.06,1.64) | 0.012 |
| *Depression symptoms | 1.09 | (1.06,1.12) | <0.001 |
| *Fall down | 1.47 | (1.02,2.15) | 0.041 |
| *Cognitive function | 0.89 | (0.85,0.93) | <0.001 |
| *Stroke | 2.55 | (1.55,4.34) | <0.001 |

Data are presented as median (interquartile range [IQR]).
Number of CMM: number of comorbidities in patients with CMM
\*was represented the meaning of P-value < 0.01 or P-value < 0.05

Gender, smoking-related indices, CVAI, TyG, and TyG-BMI were not statistically significant (P>0.05).

After LASSO regression, multivariate logistic regression identified stroke (OR=2.55, 95% CI: 1.55-4.34, P<0.001) and fall down (OR=1.47, 95% CI: 1.02-2.15, P=0.041) as independent risk factors, with stroke being the strongest, while higher cognitive function (OR=0.89, 95% CI: 0.85-0.93, P<0.001) was a protective factor.

#### Nomogram predictor selection

From the 16 variables that showed statistical significance in univariate logistic analysis, we performed LASSO regression with 10-fold cross-validation. As the regularization parameter λ increased, the regression coefficients shrank toward zero, reducing the number of predictors with non-zero coefficients (Figure 2A). The optimal λ was determined as one standard error above the minimum (λ = 0.0286, log λ = −3.5543), rather than the minimum λ value (λ = 0.0059, log λ = −5.1378) (Figure 2B). This process identified nine predictors with non-zero coefficients: depression symptoms, cognitive function, stroke, maximum left-hand grip strength, maximum right-hand grip strength, number of CMM, age, fall down, and social activity. These nine variables were then entered into multivariate logistic regression, which ultimately retained six independent predictors (all P < 0.05) for the final nomogram (Table 3).

**Figure 2.**
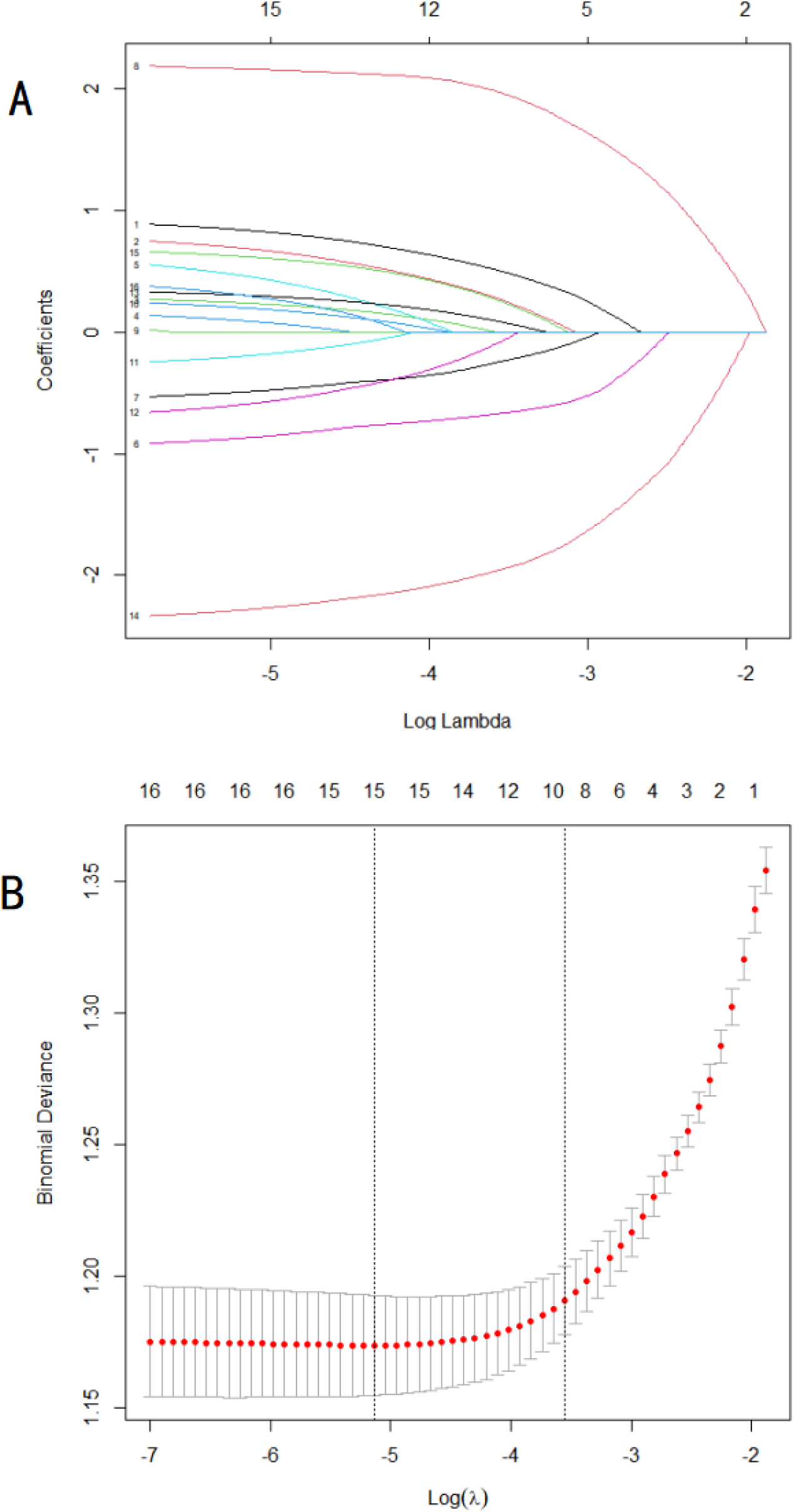
Optimize the screening variables by the LASSO regression.(A) LASSO coefficient path diagram.(B) LASSO cross validation curve to identify the optimal lambda through 10-fold cross-validation.

### Development of the prediction model (nomogram)

The final nomogram incorporating the selected predictors is shown in Figure 3A. The risk of disability in elderly CMM patients is determined by summing the allocated points for each predictor; a higher total score indicates greater risk. A dynamic nomogram is also available online at https://xjbsashjtdx.shinyapps.io/DynamicNomogram/ for clinical use, where entering the predictor values yields the predicted disability probability (Figure 3B).

**Figure 3A.**
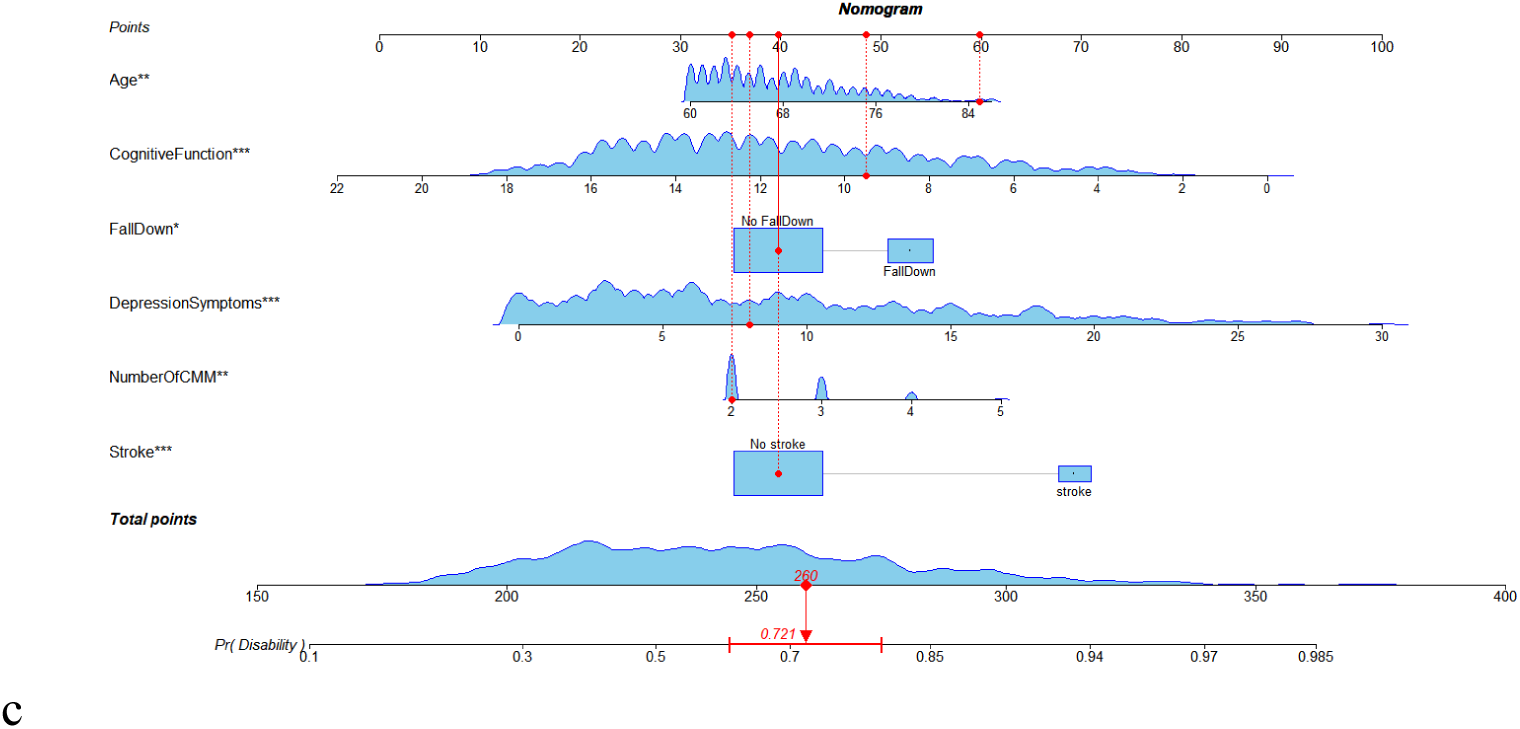
Nomogram for predicting the risk of disability in elderly patients with CMM

**Figure 3B.**
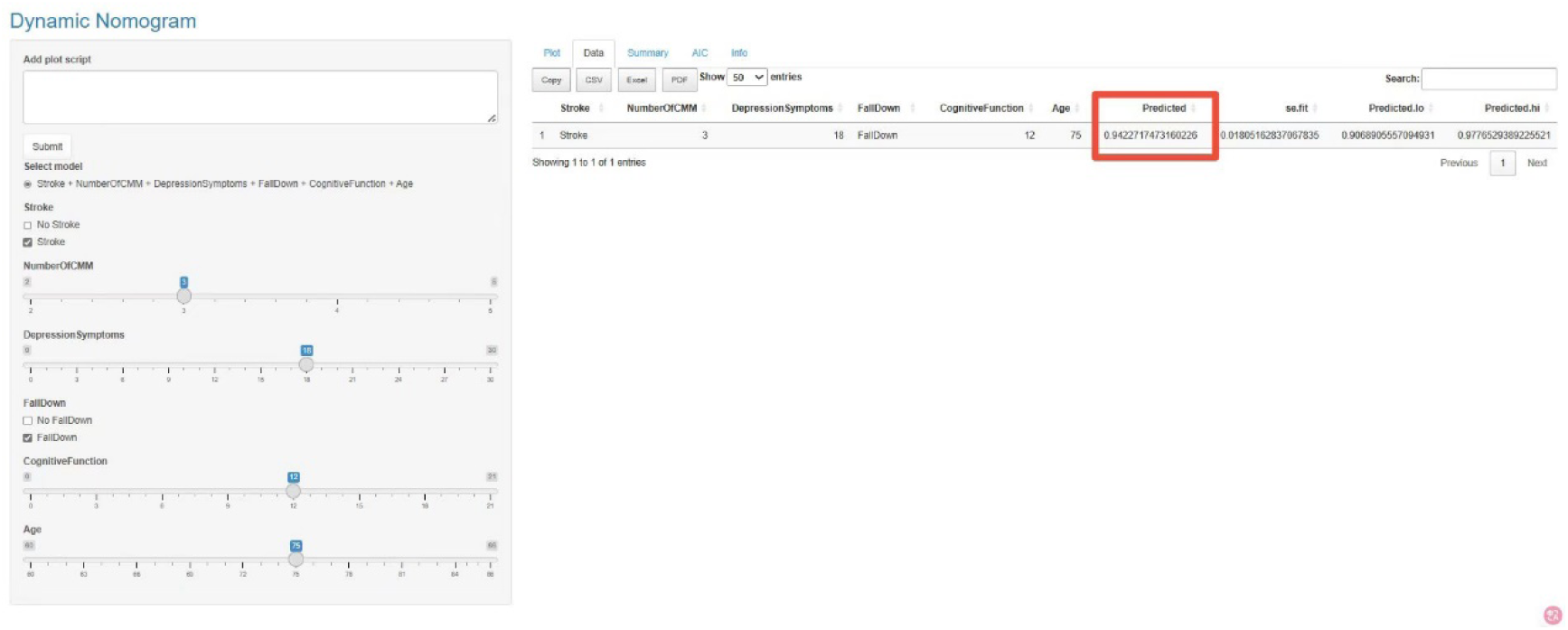
Dynamic Nomogram for predicting the risk of disability in elderly patients with CMM

### Validation and evaluation of the nomogram

The discriminative performance of the nomogram model was evaluated using receiver operating characteristic(ROC) curves. The AUC for the nomogram model were 0.746 (95% CI: 0.715–0.776) for the training set (Figure 4A), 0.740 (95% CI: 0.693–0.787) for the internal validation set (Figure 4B), and 0.754 (95% CI: 0.737–0.769) for the external validation set (Figure 4C). These results suggest that the nomogram model possesses robust discriminative ability and predictive power, effectively distinguishing between patients with and without disability.

**Figure 4.**
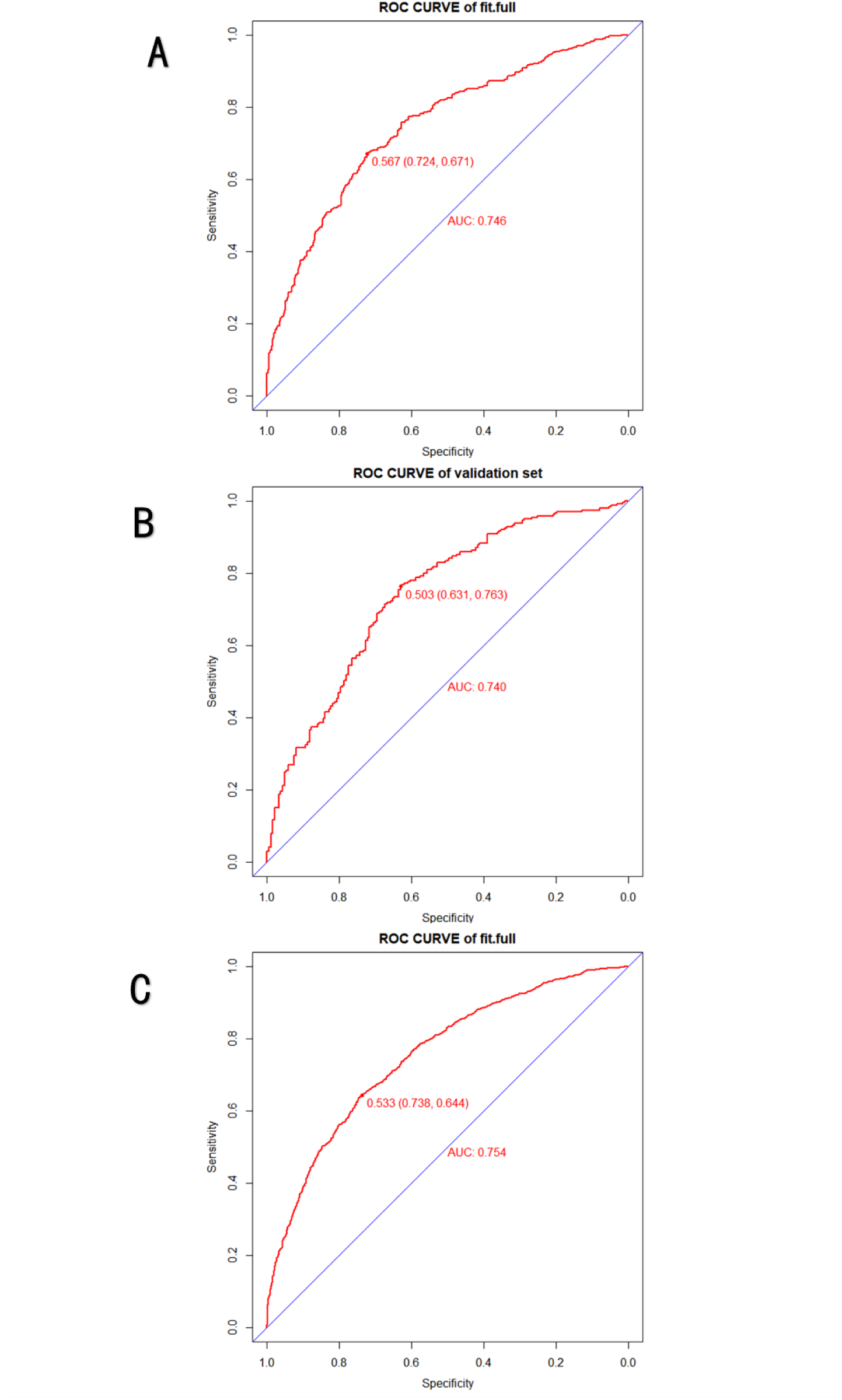
(A)ROC curve of nomogram prediction model derived from the training dataset. (B)ROC curve of nomogram prediction model derived from the internal validation dataset. (C)ROC curve of nomogram prediction model derived from the external validation dataset.

The model’s calibration was evaluated using the Hosmer-Lemeshow goodness-of-fit test and calibration curves. The P-values for the training (Figure 5A), internal validation (Figure 5B), and external validation sets (Figure 5C) were 0.9273, 0.373, and 0.7675, respectively (all P-values were > 0.05), while the Brier scores were 0.199, 0.204, and 0.201, respectively (all Brier scores were < 0.25). The results demonstrate that the model exhibits excellent calibration, indicating a high degree of concordance between the predicted and actual probabilities of disability.

**Figure 5.**
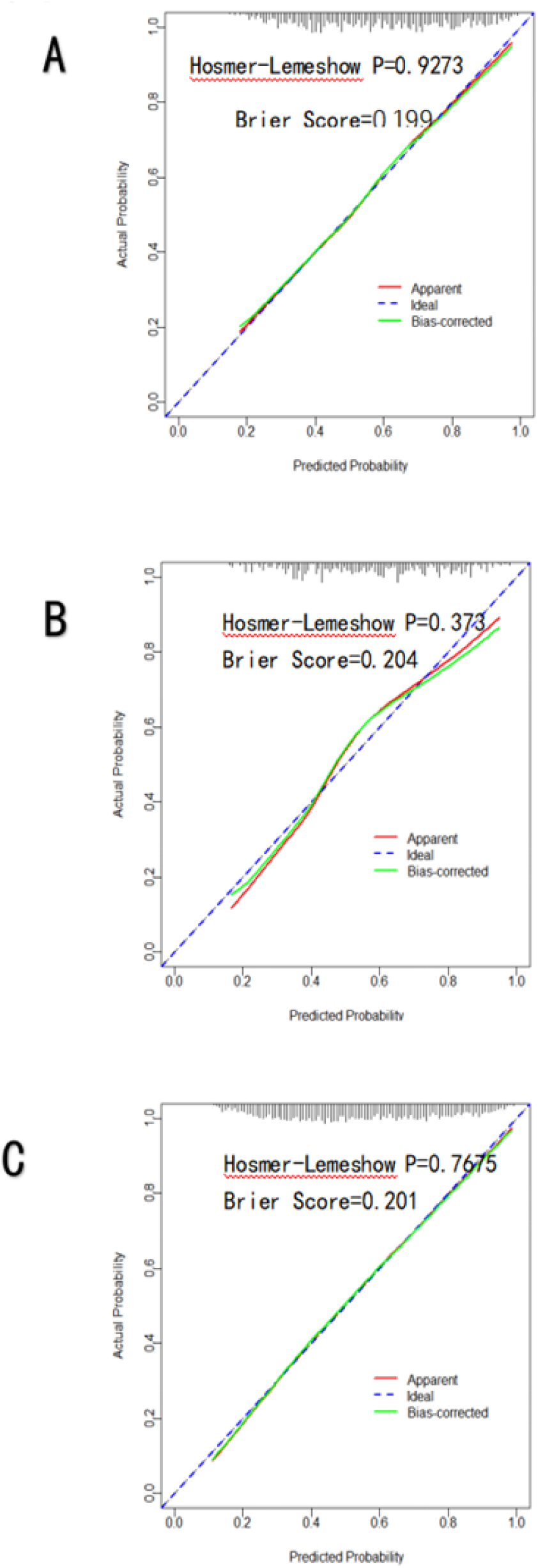
(A)Calibration curve of nomogram prediction model derived from training dataset.(B)Calibration curve of nomogram prediction model derived from internal validation dataset. (C) Calibration curve of nomogram prediction model derived from external validation dataset.

The clinical validity of the model was evaluated using DCA. The DCA indicated that the net benefit of the predictive model across the training set (Figure 6A), internal validation set (Figure 6B), and external validation set (Figure 6C) was significantly greater than that of the 2 extreme scenarios, thereby highlighting the model’s significant clinical utility.

**Figure 6.**
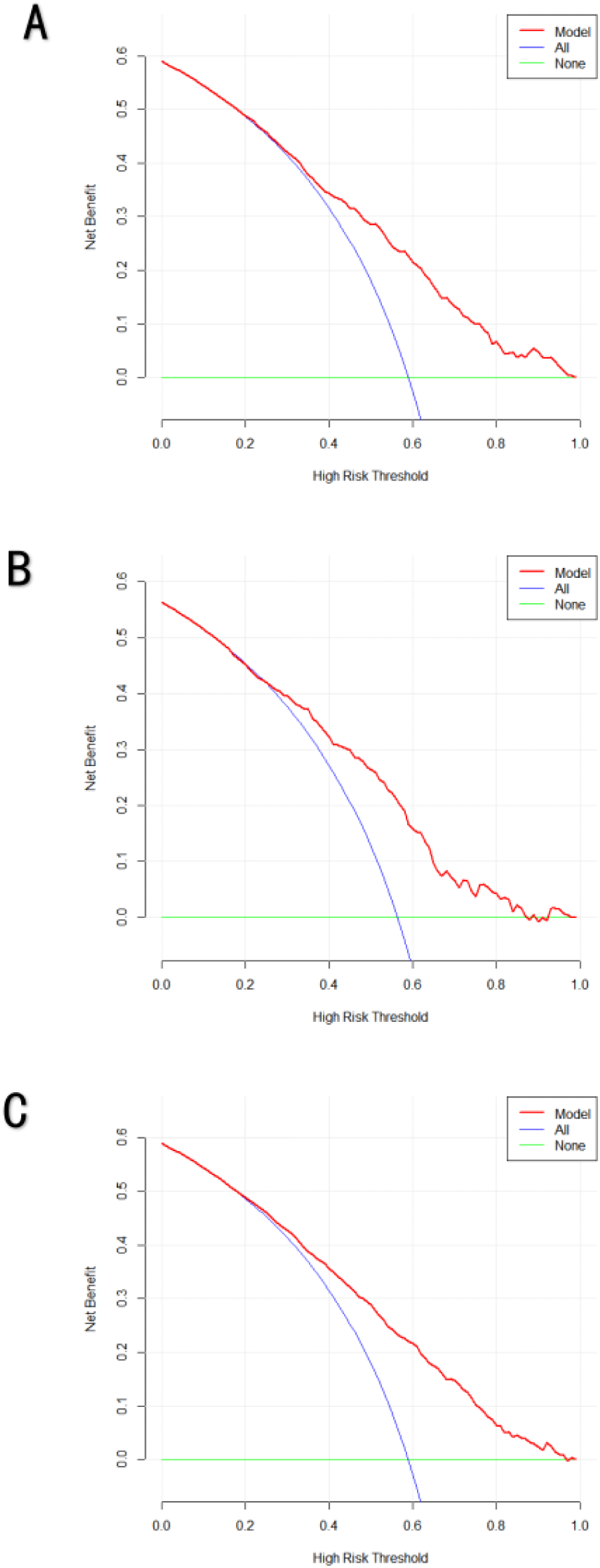
(A)DCA analysis of nomogram prediction model derived from for the training dataset.(B)DCA analysis of nomogram prediction model derived from the internal validation dataset. (C)DCA analysis of nomogram prediction model derived from the external validation dataset.

### Mediation analysis results

Mediation analysis was conducted to further explore the associations between Number of CMM, depression symptoms, cognitive function, stroke, fall down, and disability.

When Number of CMM was set as the independent variable, no variable showed a mediating effect. When depression symptoms served as the independent variable, Maximum Right-hand grip strength, Maximum Left-hand grip strength, and Fall down exhibited partial positive mediating effects, with the proportions of mediating effects being 3.4%, 5.2%, and 2.9%, respectively(Figure 7A). When cognitive function was the independent variable, Maximum Right-hand grip strength, Maximum Left-hand grip strength, Average walking speed, and Fall down demonstrated partial negative mediating effects, accounting for 6.5%, 5.9%, 6.3%, and 2.4% of the total effects, respectively(Figure 7B). When Stroke was the independent variable, Maximum Left-hand grip strength and Average walking speed showed partial positive mediating effects, with mediating effect ratios of 12.3% and 6.7%, respectively(Figure 7C). Finally, when Fall down was set as the independent variable, no variable exhibited a mediating effect.(Supplementary Table1-15)

**Figure 7A.**
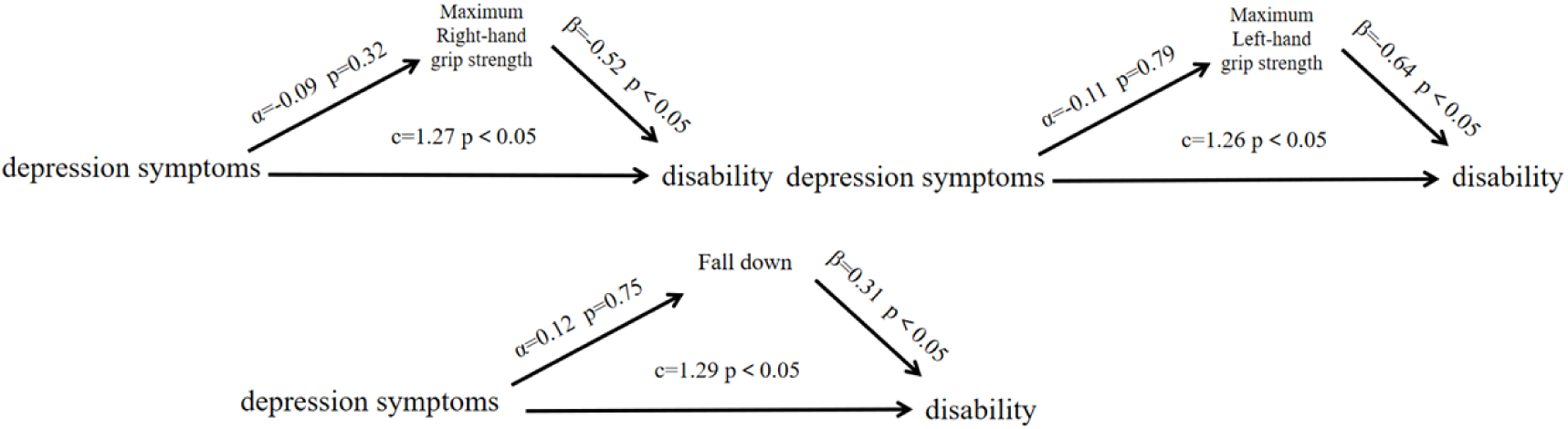
Analysis of the mediation between depression symptoms and disability

**Figure 7B.**
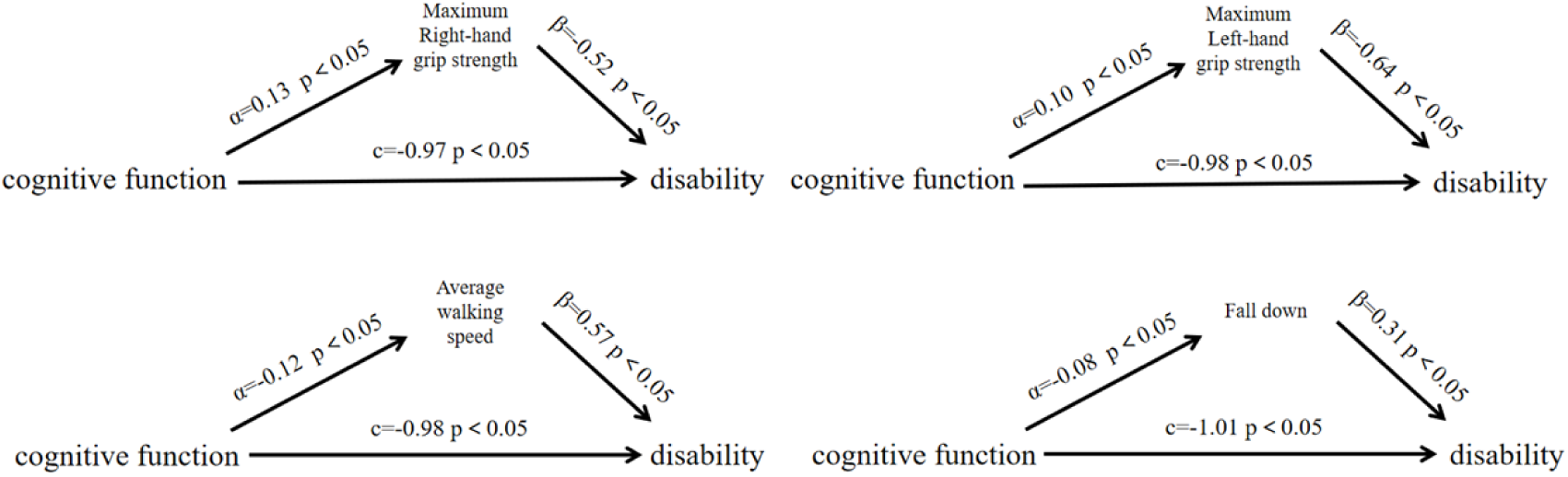
Analysis of the mediation between cognitive function and disability

**Figure 7C.**
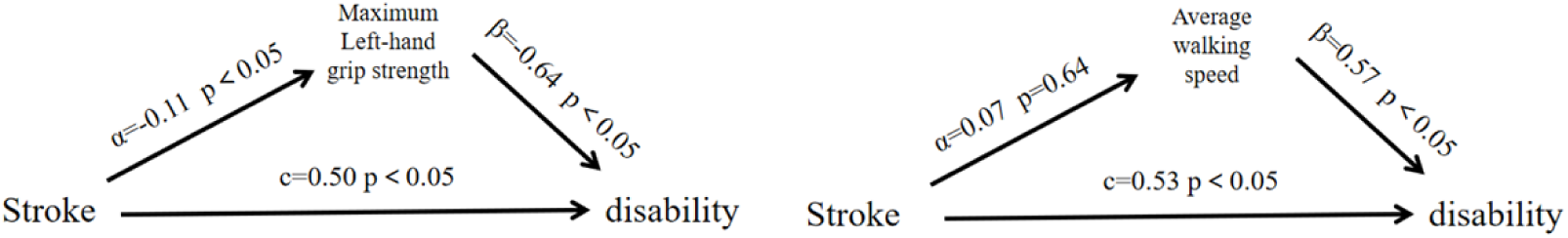
Analysis of the mediation between Stroke and disability

### Results of RCS analysis and threshold effect analysis

RCS analysis revealed an inverted U-shaped nonlinear association between cognitive function and disability. Threshold effect analysis identified an inflection point at 12.043, with an OR of 0.84 (95% CI: 0.81–0.87), and The likelihood ratio test was significant (p = 0.0007)(Figure 8 and Table 4).Whereas depression symptoms, fall down, and stroke showed linear associations(Supplementary Figure1-3 and table16).

**Figure 8.**
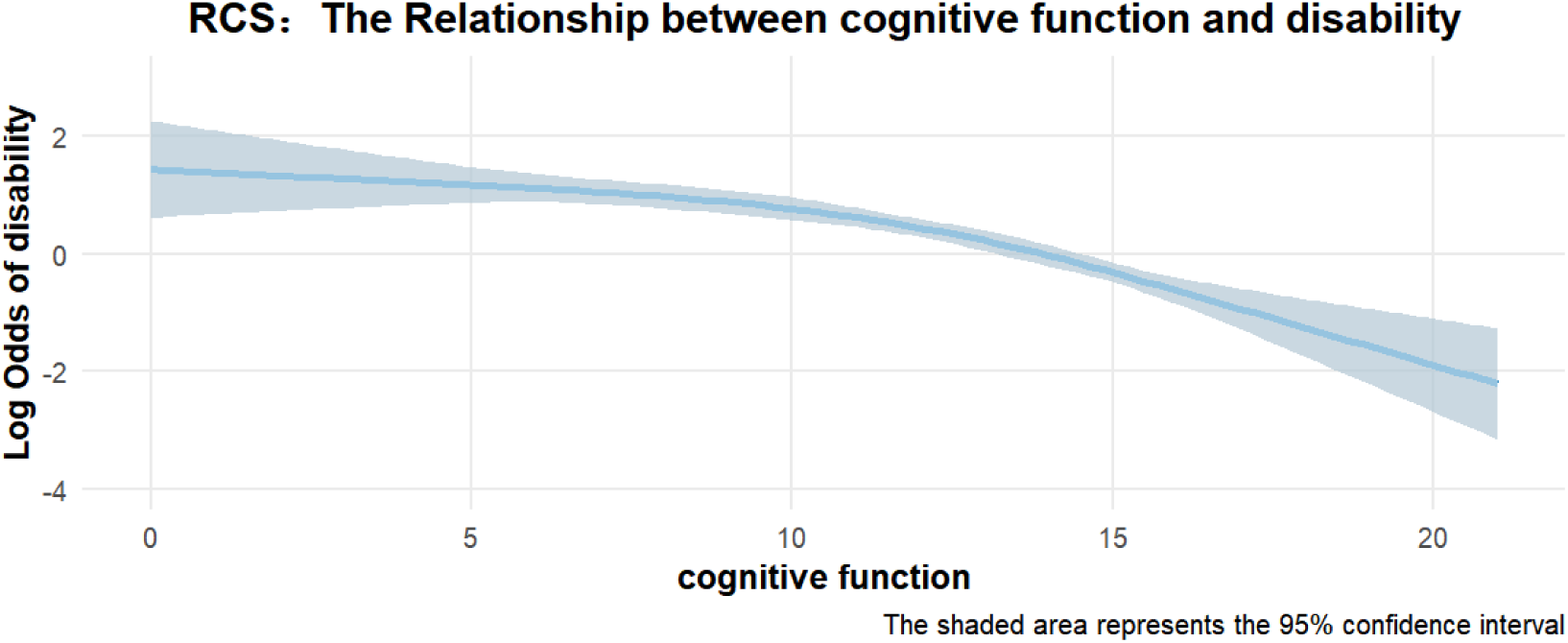
RCS curve of cognitive function

**Table 4.** Threshold Analysis result of cognitive function.

| Disability | OR(95%CI),P value |
| --- | --- |
| Model 1: Standard linear regression fit. | 0.84 ( 0.81-0.87 ) , < 0.001 |
| Model 2: Two-segment linear regression fit |  |
| Inflection point ( cognitive function ) | 12.043 |
| < 12.043 | 0.91 ( 0.86-0.96 ) , < 0.001 |
| > 12.043 | 0.74 ( 0.68-0.80 ) , < 0.001 |
| Inflection point and likelihood ratio P-value | 0.0007 ( < 0.05 ) |

### Linear dose-response relationship between number of CMM and disability with a plateau

Among the participants, the distribution of Number of CMM was: CMM=2 (n=876), CMM=3 (n=410), CMM=4 (n=125), CMM=5 (n=13). Compared with CMM=2, CMM=3 (OR=1.40, 95% CI: 1.10–1.78, P=0.01) and CMM=4 (OR=1.72, 95% CI: 1.17–2.58, P=0.01) showed significantly higher odds of disability. CMM=5 had a large point estimate (OR=10.08) but a very wide confidence interval (95% CI: 1.97–184.12, P=0.03). After Tukey adjustment, only the differences between CMM=2 and CMM=3 (logOR=−0.334, P=0.032) and between CMM=2 and CMM=4 (logOR=−0.543, adjusted P=0.036) remained significant; no other pairwise comparisons were significant.The results showed that the CMM≥4 group had an OR of 1.92 (95% CI: 1.31–2.85, P = 0.001). After Tukey’s correction, the difference between CMM=2 and CMM≥4 remained significant (logOR = –0.653, P = 0.0027), whereas the difference between CMM=3 and CMM≥4 was not significant.

To further assess the linear dose-response relationship, three trend analyses were performed. A likelihood-ratio test showed that a linear model fitted the data as well as the factor model (P=0.258). Orthogonal polynomial contrasts revealed a significant linear trend (P=0.023), with non-significant quadratic and cubic trends. The Cochran-Armitage test confirmed a strong linear increase in disability proportion across CMM levels (P<0.001). Collectively, these results support a linear dose-response relationship between higher Number of CMM and increased disability risk(Supplementary Table 17-21).

## Machine learning results

### Selection of characteristic variable

The Boruta algorithm identified 22 candidate predictors from the 46 initial variables, including Confirmed and Tentative variables( Figure 9 and Supplementary Table 22). When LASSO regression was applied directly to the same 46 variables, nine predictors with non-zero coefficients were obtained(Supplementary Figure 4-5 and Table 23).

**Fig 9.**
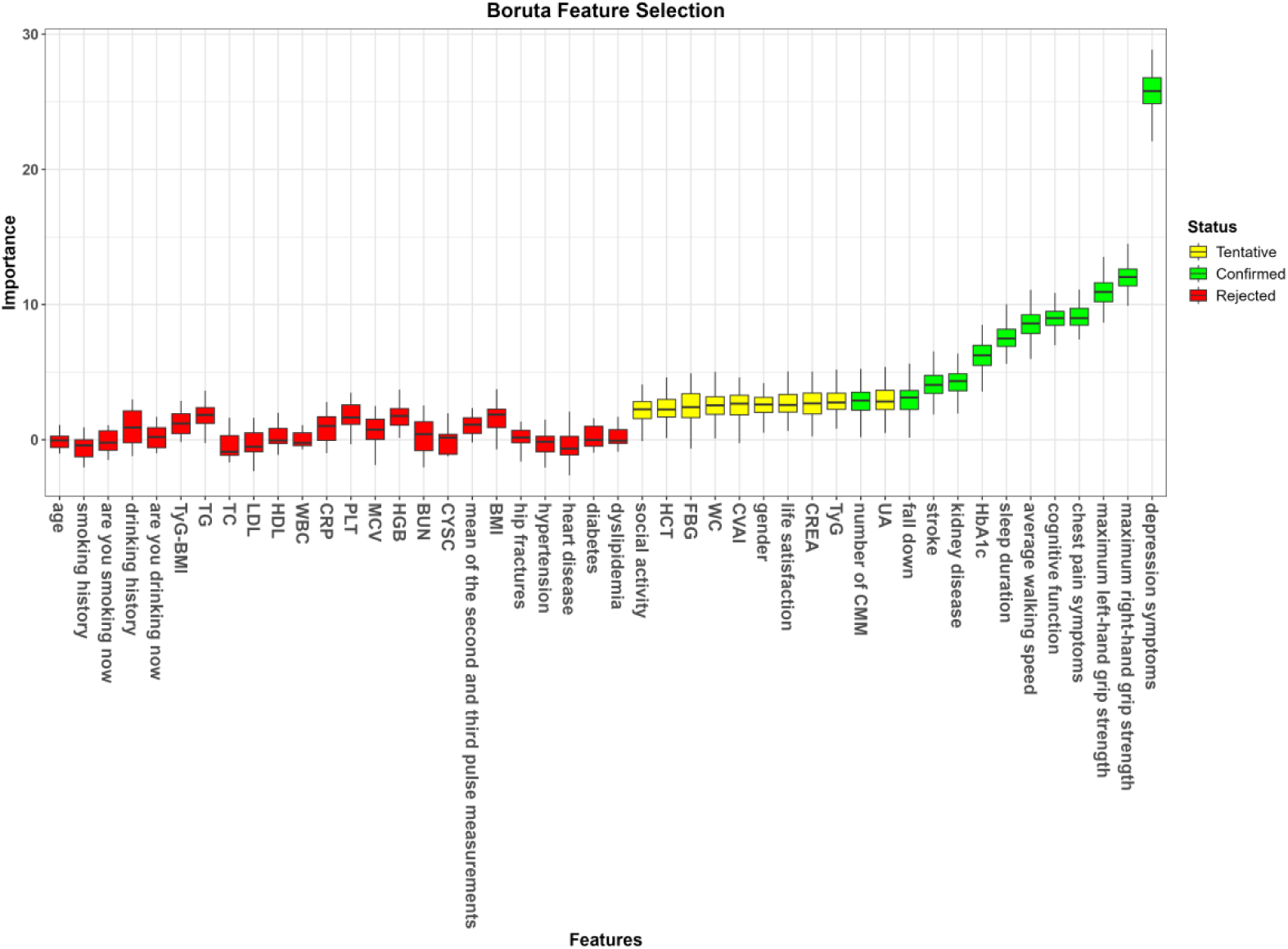
Boruta Feature Selection

Notably, these nine variables constituted exactly the intersection of the two methods(Supplementary Table 24). They were: kidney disease, stroke, chest pain symptoms, average walking speed, depression symptoms, fall down, maximum left-hand grip strength, maximum right-hand grip strength, and cognitive function. These indicators covered mental state, physical function, metabolic indicators, and chronic disease history, forming the final predictor system for disease risk prediction.

### Model performance comparisons

Five machine learning models were established for disease risk prediction, including LR, RF, SVM, LGB, and XGBoost. In the training cohort, the AUC values were 0.796 for LR, 0.737 for RF, 0.755 for SVM, 0.716 for LGB, and 0.716 for XGBoost(Supplementary Table 25). In the internal validation cohort, the RF model demonstrated superior predictive performance, with an AUC of 0.782, accuracy of 0.732, sensitivity of 0.640, specificity of 0.777, and the lowest Brier score of 0.1736(Table 5), followed by LR (AUC = 0.776) and SVM (AUC = 0.760)(Figure 10A), while LGB and XGBoost achieved AUC values of 0.716 and 0.721, respectively(Figure 10B). Calibration curves showed that the predicted probabilities of the RF model were highly consistent with the actual outcomes(Figure 10C and Figure 10D).DCA confirmed that the RF model provided the highest and most stable net benefit across a wide range of threshold probabilities, indicating favorable clinical practicability(Figure 10E and Figure 10F). Based on the above results, the RF model was selected as the optimal prediction model for subsequent analyses.

**Figure 10A.**
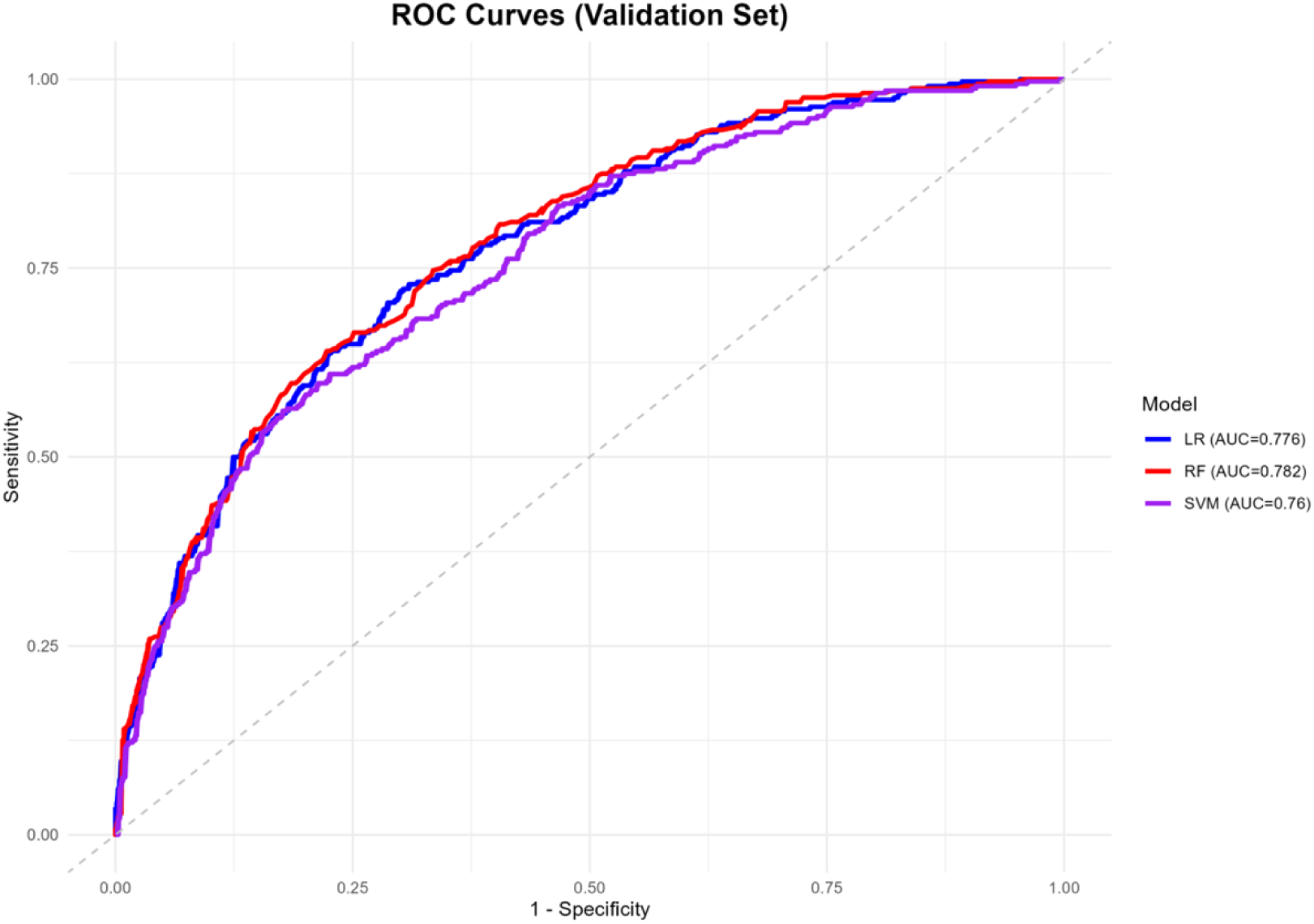
ROC Curves of LR, RF and SVM Models in the Validation Set

**Figure 10B.**
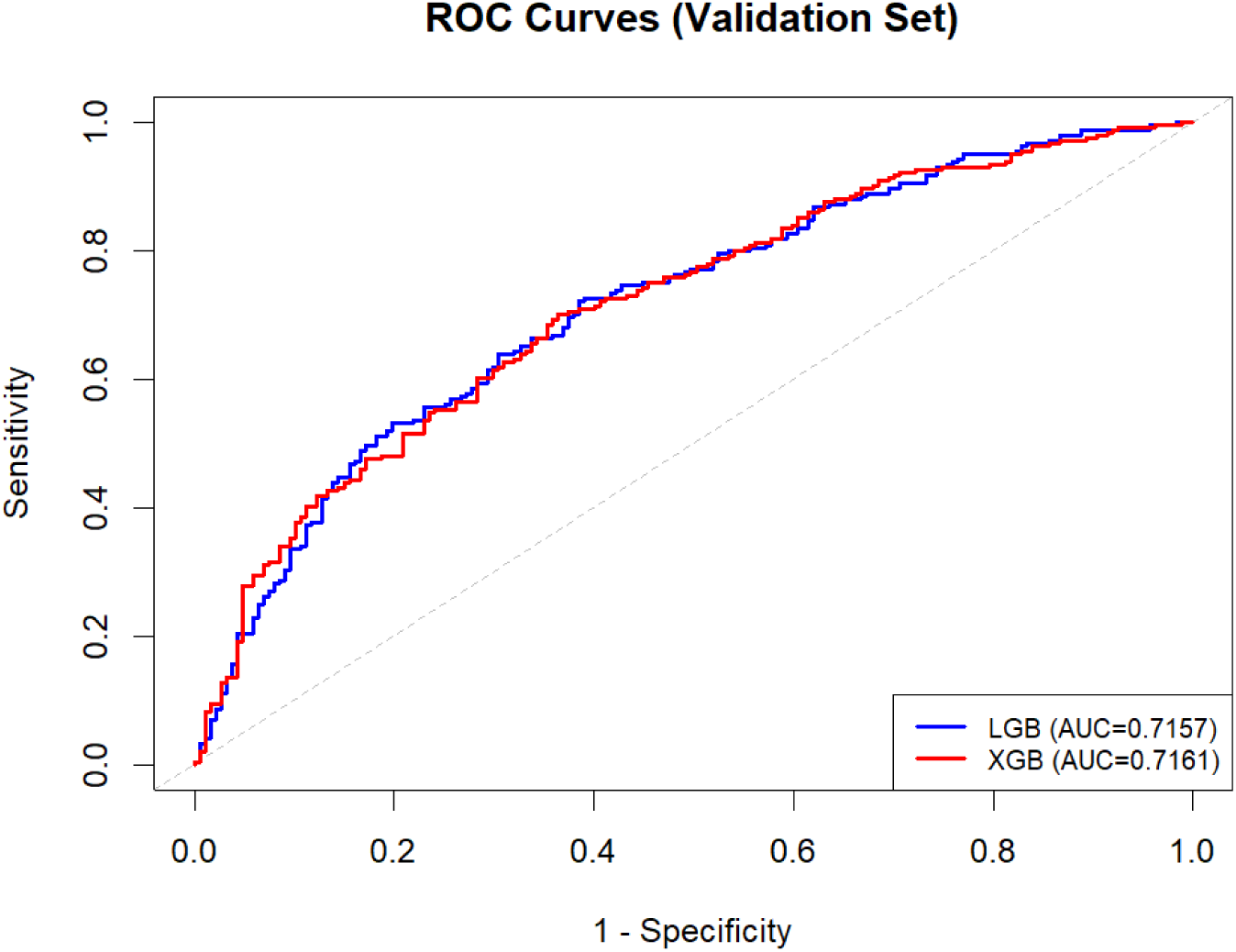
ROC Curves of LGB and XGB Models in the Validation Set.

**Figure 10C.**
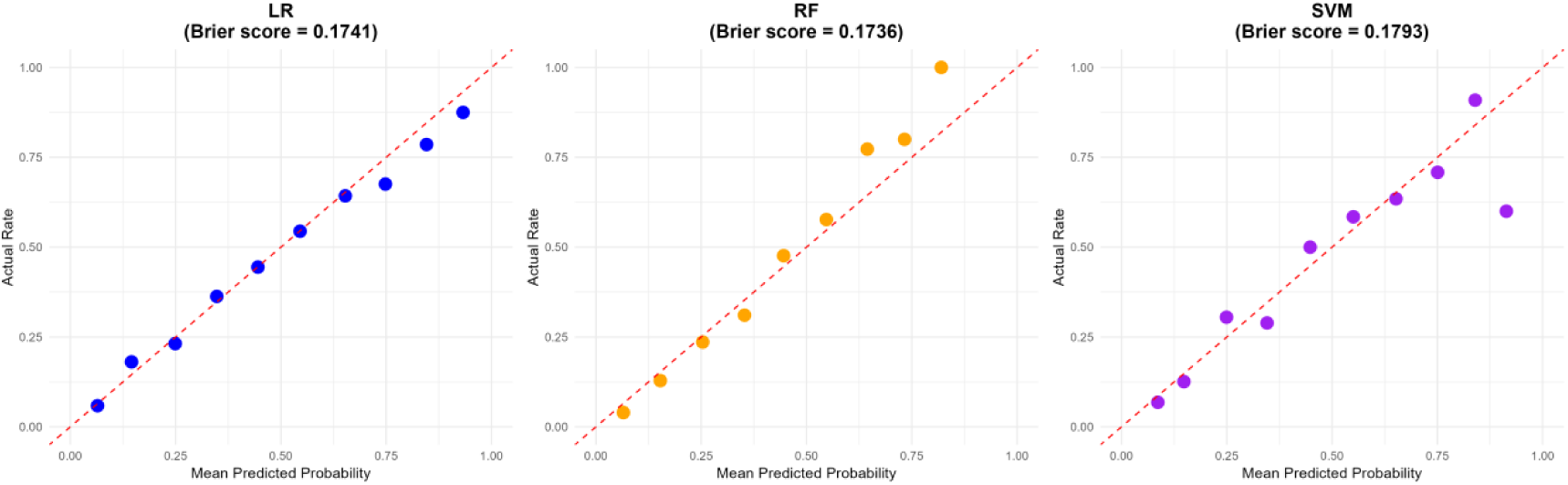
Calibration Curves of LR, RF and SVM Models.Calibration curves of the LR, RF and SVM models.

**Figure 10D.**
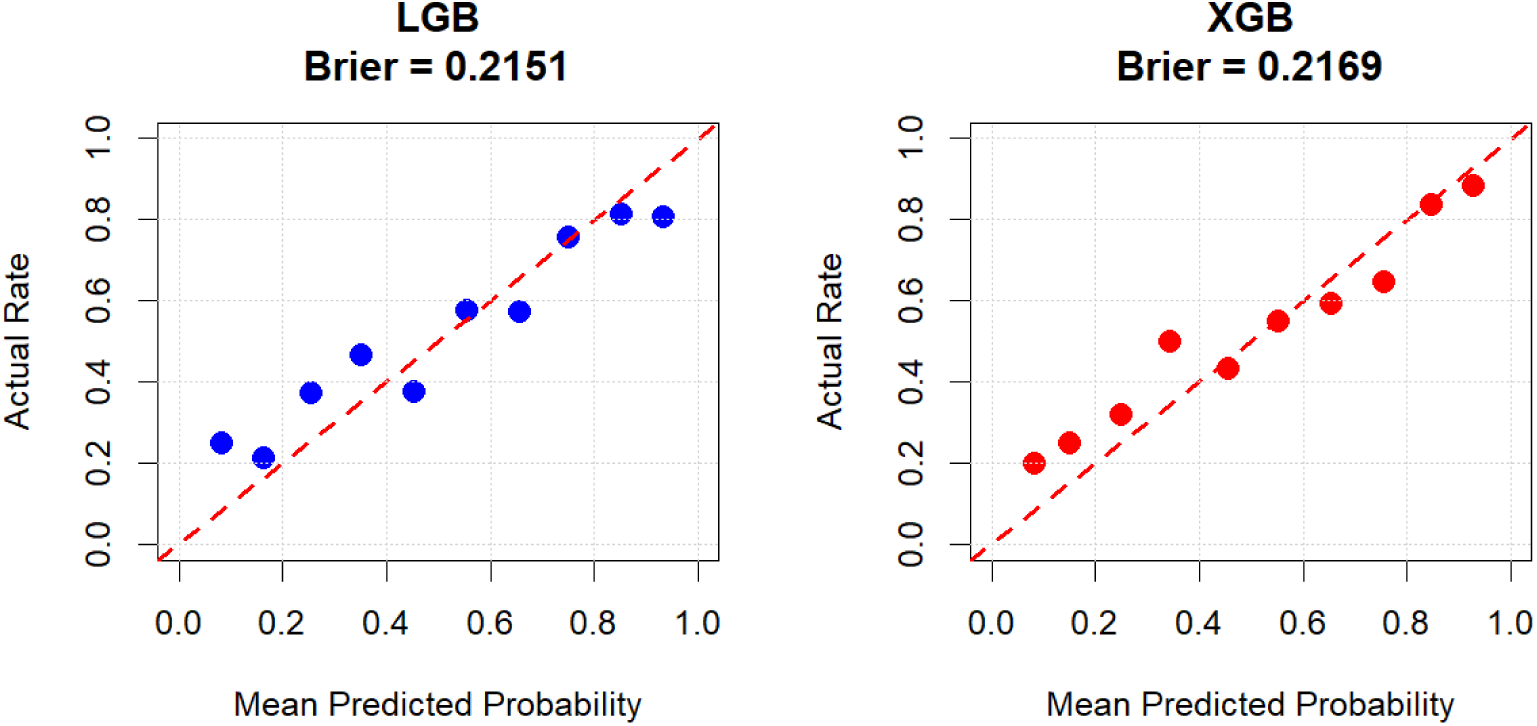
Calibration Curves of LGB and XGB Models.

**Figure 10E.**
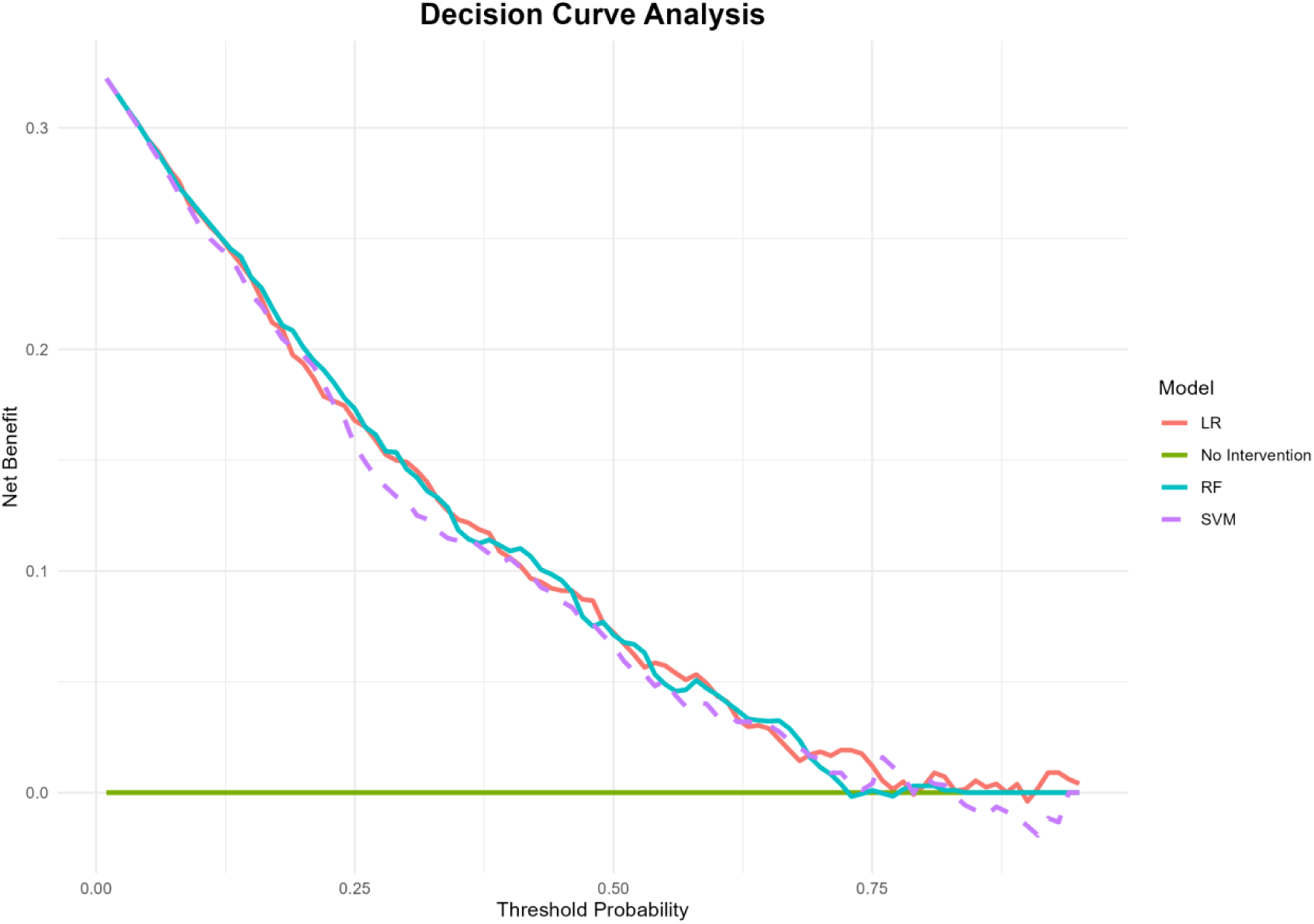
Decision Curve Analysis of LR, RF and SVM Models.

**Figure 10F.**
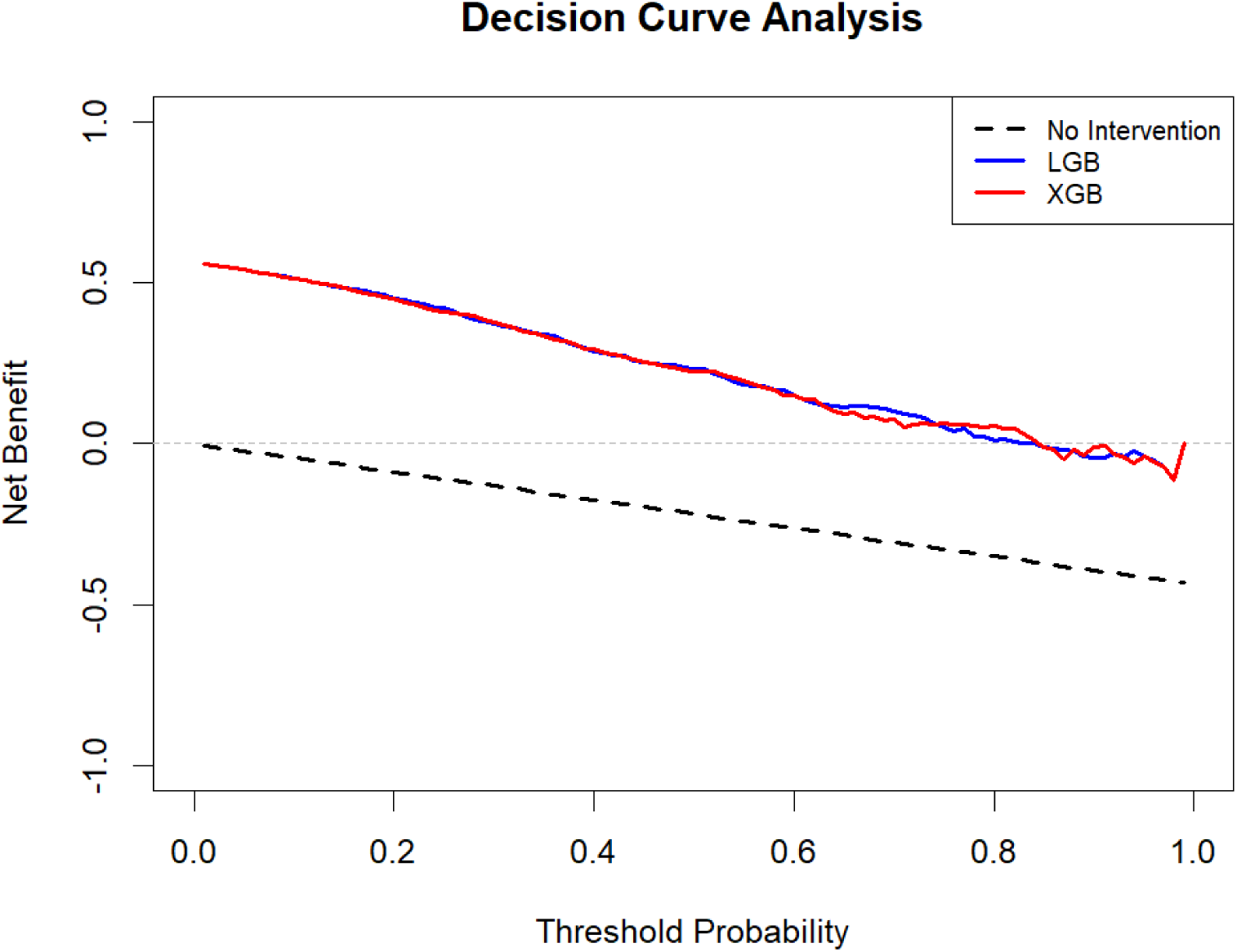
Decision Curve Analysis of LGB and XGB Models.

**Table 5:** Performance Comparison of Five Machine Learning Models on Validation Set.

| Model | AUC | Accuracy | Sensitivity | Specificity | Precision | Recall | F1 |
| --- | --- | --- | --- | --- | --- | --- | --- |
| LGB | 0.7157 | 0.6682 | 0.7095 | 0.615 | 0.7037 | 0.7095 | 0.7066 |
| XGB | 0.7212 | 0.6776 | 0.7054 | 0.6417 | 0.7173 | 0.7054 | 0.7113 |
| LR | 0.776 | 0.703 | 0.729 | 0.691 | 0.536 | 0.729 | 0.618 |
| RF | 0.782 | 0.732 | 0.640 | 0.777 | 0.585 | 0.640 | 0.611 |
| SVM | 0.760 | 0.737 | 0.561 | 0.824 | 0.609 | 0.561 | 0.584 |

### Model explanation

To interpret the RF model, we employed SHAP at both global and individual levels. Globally, feature importance was ranked by mean absolute SHAP values. The top three predictors were depression symptoms, average walking speed, and maximum right-hand grip strength. Additional important predictors included cognitive function, maximum left-hand grip strength, sleep duration, and metabolic markers (HbA1c, CREA, UA, HCT, FBG, TyG, CVAI; Supplementary Figure 7). Notably, kidney disease had higher predictive value than number of CMM and stroke.The dot plot (Supplementary Figure 6) further revealed that higher levels of HbA1c, HCT, FBG, TyG, and CVAI were associated with increased risk.

At the individual level, the SHAP waterfall plot and force plot(Supplementary Figure 8-9) illustrated the additive contribution of each feature to the predicted probability for a single sample. Although local deviations were observed in the specific example, they did not contradict the global findings. Overall, the population-level directions were consistent with clinical knowledge and highly aligned with the results from traditional analyses. Thus, the SHAP analysis provides good interpretability for the machine learning model.

## Discussion

CMM remains a major public health burden in the elderly, characterized by high morbidity, disability, mortality, and healthcare costs [16–18]. In this study, we developed a dynamic nomogram integrating six predictors (depression symptoms, cognitive function, stroke, number of CMM, age, and fall down) to predict disability risk in elderly CMM patients using CHARLS. The model demonstrated good discrimination, calibration, and clinical applicability. To our knowledge, this is the first practical dynamic nomogram for this population. By integrating traditional regression, mediation analysis, RCS with threshold analysis, linear trend assessment for number of CMM (levels 2–5) and machine learning, we further addressed three key questions: (1) whether Metabolic indicators predict disability; (2) whether depression symptoms (mental health) outweigh grip strength and walking speed (physical function); and (3) whether the number of CMM exhibits a plateau rather than a purely linear trend.

Regarding the first question, in traditional logistic regression, metabolic markers were not significant. However, in the RF model with SHAP, they emerged as important predictors, with higher levels of HbA1c, HCT, FBG, TyG, and CVAI associated with increased disability risk. We categorized these markers into glucose metabolism (HbA1c, FBG), lipid-obesity (TyG, CVAI), renal function (CREA, UA), and hematological (HCT). Why the discrepancy? Traditional logistic regression assumes linear, additive relationships and is sensitive to multicollinearity and sample size, whereas RF captures nonlinear interactions typical of metabolic dysregulation.

Thus, inexpensive routine blood markers can be clinically relevant even when conventional models miss them. A notable contrast also emerged for chronic diseases. In traditional regression, stroke was the strongest predictor (OR = 2.55). In contrast, the RF model ranked kidney disease higher than both stroke and number of CMM, suggesting that kidney disease captures complex, nonlinear interactions in multimorbidity. These contrasting results demonstrate that machine learning, especially RF with SHAP, can reveal hidden patterns beyond traditional regression.

Regarding the second question, Depression symptoms consistently outperformed grip strength and walking speed in both traditional and machine learning models, highlighting the dominant role of psychological status over physical function in disability risk.Mediation analysis further elucidated the pathways. Grip strength and falls partially mediated the effects of depression symptoms and cognitive function, while walking speed mediated the effects of cognitive function and stroke. Notably, left-hand grip strength was the strongest mediator, accounting for 12.3% of the effect of stroke on disability. Thus, functional decline (reduced grip strength, slower walking speed) and falls are not only independent risk factors but also intermediate mechanisms linking depression, cognitive impairment, and stroke to disability.

Regarding the third question, the number of CMM showed a positive dose-response relationship with a plateau after CMM=3, rather than a purely linear trend. After Tukey adjustment, CMM=3 and CMM=4 remained significantly associated with higher disability risk than CMM=2; CMM=5 gave an unstable OR (10.08). Sensitivity analysis merging CMM=4 and CMM=5 into CMM≥4 yielded a significant difference from CMM=2, but not from CMM=3, confirming a plateau.Trend analyses showed a significant linear component.

Our findings largely agree with prior studies. Lee et al. found that cancer, diabetes, hypertension, stroke, heart failure, and lung diseases were associated with disability [44]. Zhou et al. reported that older age, higher BMI, and hypertension increased ADL disability risk [45]. Zhang et al. included age, education, gait speed, cognitive function, and depression symptoms to predict IADL disability [46]. Han et al. identified right-hand grip strength and depression symptoms as top predictors [23]. Our study, conducted in elderly CMM patients, largely agrees with these findings and extends them by uncovering a plateau in the number of CMM-disability relationship and highlighting the importance of kidney disease over stroke in multimorbidity. In traditional analysis, stroke was the strongest predictor of disability (OR = 2.55), followed by falls (OR = 1.47). Stroke is a leading cause of disability worldwide [47], and falls are associated with long-lasting ADL disability and cognitive decline [48, 49]. Cognitive impairment further exacerbates post-stroke functional decline and recurrence risk [50, 51].

Importantly, unlike most previous studies, we incorporated a wide range of laboratory variables. SHAP analysis from the RF model showed that higher levels of HbA1c, CREA, UA, HCT, FBG, TyG, and CVAI were associated with increased disability risk. Mechanistically, elevated HbA1c is linked to cardiovascular events and mortality [52]. CREA reflects not only renal function but also muscle metabolism and physiological vulnerability, contributing to CMM pathophysiology through impaired muscle function and inflammatory homeostasis[53]. UA activates NADPH oxidase and aggravates oxidative stress while inhibiting endothelial nitric oxide synthesis, promoting vasoconstriction and renal dysfunction, thereby facilitating CMM progression[54]. Elevated HCT is associated with reduced reperfusion in acute stroke [55]. Elevated FBG under chronic hyperglycemia increases reactive oxygen species and inflammatory cytokines, inducing endothelial dysfunction and pancreatic β-cell damage, driving the pathological continuum from dysglycemia to cardiometabolic disorders[56]. Additionally, TyG is associated with new-onset cardiovascular disease [33], and CVAI predicts metabolic disorders and CVD events [36–39].

This study has several strengths, including a broad disability definition, a clinically applicable dynamic nomogram with good performance, integration of traditional and machine learning methods, predictive value of Metabolic indicators despite their non-significance in conventional models, and identification of a plateau in the num ber of CMM-disability relationship. However, several limitations should be acknowledged. First, routine blood metabolic markers were non-significant in traditional analyses, likely due to sample size and selection bias, warranting multi-center studies. Second, CHARLS data require external validation in other populations. Third, the retrospective design without follow-up precludes causal inference.

In conclusion, using six predictors, we developed a dynamic nomogram to predict disability in elderly CMM patients. Integrated analyses revealed that routine blood metabolic markers predict disability, and depression symptoms outperform physical function. We also identified a plateau in the number of CMM-disability relationship. Stroke was the strongest traditional predictor, whereas kidney disease was more important in machine learning. Our nomogram is clinically practical, and incorporating metabolic markers may enhance future models, supporting routine blood tests for early risk identification.

## Supplementary Information

### Electronic supplementary material

Below is the link to the electronic supplementary material.

## Abbreviations

WHO: The World Health Organization
CVD: Cardiovascular Disease
CMM: Cardiometabolic Multimorbidity
BMI: Body Mass Index
ADL: Activities of Daily Living
IADL: Instrumental Activities of Daily Living
CHARLS: China Health and Retirement Longitudinal Study
TyG: Triglyceride Glucose Index
TyG-BMI: Triglyceride Glucose-Body Mass Index
CVAI: Chinese Visceral Adiposity Index
IR: Insulin Resistance
HbA1c: Glycated Hemoglobin A1c
FBG: Fasting Blood Glucose
TG: Triglycerides
TC: Total Cholesterol
LDL: Low-Density Lipoprotein Cholesterol
HDL: High-Density Lipoprotein Cholesterol
WBC: White Blood Cells
CRP: C-Reactive Protein
PLT: Platelets
MCV: Mean Corpuscular Volume
HGB: Hemoglobin
HCT: Hematocrit
CREA: Creatinine
BUN: Urea Nitrogen
CYSC: Cystatin C
UA: Uric Acid
WC: Waist Circumference
CES-D-10: Center for Epidemiological Studies Depression Scale
OR: Odds Ratio
CI: Confidence Interval
LASSO: Least Absolute Shrinkage and Selection Operator
AUC: Area Under the ROC curve
DCA: Decision Curve Analysis
RCS: restricted cubic spline
LR: Logistic Regression
RF: Random Forest
SVM: Support Vector Machine
LGB: Light Gradient Boosting
XGBoost: Xtreme Gradient Boosting
SHAP: SHapley Additive exPlanation
ROC: Receiver Operating Characteristic

## Acknowledgement

We would like to thank all the participants of this project and investigators for collecting the data.

## Author contributions

HX performed experiments, analyzed data, and wrote the main manuscript text. ZX carried out experiments and contributed to data analysis. YS supervised the research and provided guidance on the study design.SS oversaw the research, offered study design guidance, and reviewed the manuscript. Machine learning and ST both provided technical support and conducted experiments. LJ, XH, ZS, YW, and CS all contributed to writing and revising the manuscript.

## Data Availability

No original data were produced for this research. All underlying individual-level data come from the CHARLS database, which can only be obtained by submitting an official application at http://charls.pku.edu.cn/en.

## Ethical approval

Ethics approval for the study was granted by the Ethics Review Committee of Peking University, and all the participants provided signed informed consent at the time of participation.The study methodology was carried out following approved guidelines.

## Consent for publication

All authors gave consent for the publication of the article.We confirm that all authors have participated in the work and have reviewed and approved the final version of the manuscript. This manuscript is original, has not been published previously, and is not under consideration for publication by any other journal. We affirm that no part of the text has been plagiarized, and all sources are appropriately cited. Furthermore, we understand that it is our responsibility to obtain permission for any reproduced figures or tables and to cover any associated fees, with such permissions to be secured before the manuscript’s final acceptance.

